# Early-childhood temperament deviations mark psychiatric risk into early adulthood

**DOI:** 10.64898/2026.03.27.26349492

**Authors:** J. Kopal, N.R. Bakken, P. Parekh, A.A. Shadrin, P. Jaholkowski, L.A.R. Ystaas, N. Parker, O.B. Smeland, E.P. Tissink, I.E. Sønderby, K.S. O’Connell, O. Frei, A.M. Dale, O.A. Andreassen

**Affiliations:** Centre for Precision Psychiatry, Division of Mental Health and Addiction, Oslo University Hospital & Institute of Clinical Medicine, University of Oslo, Oslo, Norway; Center for Multimodal Imaging and Genetics, J. Craig Venter Institute, La Jolla, CA, USA; K.G. Jebsen Centre for Neurodevelopmental Disorders, University of Oslo and Oslo University Hospital, Oslo, Norway; Department of Medical Genetics, Oslo University Hospital & University of Oslo, Oslo, Norway; University of California San Diego, La Jolla, CA, USA; Oslo University Hospital, Oslo, Norway

## Abstract

Early-childhood temperament is associated with mental health outcomes decades later. Temperament reflects early-emerging individual differences in emotional and behavioral tendencies. These differences are relatively stable across development and shaped by both genetic and environmental influences. However, the consequences of departures from expected developmental trajectories remain largely unexplored. Using data from more than 50,000 children in the Norwegian Mother, Father and Child Cohort Study, we modeled longitudinal temperament trajectories at 1.5, 3, and 5 years of age and quantified deviations from expected development. Multivariate pattern analysis revealed latent dimensions linking these deviations to clinical diagnoses, with ADHD as the most prominent outcome. Time-to-event analysis showed that these dimensions were associated with a higher hazard of ADHD diagnosis across childhood and adolescence. Finally, genetic analyses identified loci jointly associated with temperament trajectories and ADHD, revealing age-dependent genetic effects. Together, these findings show that deviations from temperament trajectories in early childhood capture transdiagnostic vulnerability across development. Early temperament monitoring may thus serve as an indicator of later mental health risk.

## Introduction

Well before children speak their first word, they display temperamental tendencies that have long been linked to later emotional and behavioral functioning^1^. By temperament, we here refer to early-appearing individual differences in emotional and behavioral reactivity, shaped by the child’s partly biologically rooted tendency to approach and react to the environment^2^. These individual patterns are partly attributable to genetic variation^3,4^ and reflect innate predispositions that guide early behavior^5,6^. Temperament is further shaped by neurobiological maturation and environmental experiences, both of which substantially change across childhood^7–9^. Temperament therefore shows partial stability while continuing to evolve^3,10^. This combination of stability and change implies that, despite individual variability, children tend to follow broadly predictable temperament developmental trajectories^7^. When children depart from these expected trajectories, the deviations may serve as early markers of clinical vulnerability^11,12^. However, how deviations from normative trajectories in early childhood relate to later mental health outcomes remains unclear.

One of the most widely used instruments in early-childhood temperament research is the Emotionality, Activity, Shyness, and Sociability Temperament questionnaire (EAS)^13^. The EAS characterizes temperament using four core traits: *emotionality*, reflecting sensitivity and intensity of negative affect; *activity*, indexing the vigor and tempo of motor behavior; *shyness*, referring to inhibition in the presence of unfamiliar people; and *sociability*, reflecting the desire to seek and enjoy social interaction. The validity of these four traits is supported across diverse populations, ages, and measurement approaches^14–16^. Twin and family studies estimate substantial heritability for these traits, typically in the range of 30–60%^3,4^. Evidence suggests that genetic influences on temperament may vary across development^3,17^, which underscores the importance of longitudinal approaches. The EAS thus provides a well-established framework for characterizing early temperament and examining its developmental course.

Extensive longitudinal work demonstrates that early temperament predicts a wide range of later developmental, behavioral, and mental health outcomes^18–21^. These predictive links extend well into adulthood, including associations with social functioning more than three decades later ^22^. Specific psychiatric and neurodevelopmental disorders are associated with characteristic early temperamental profiles. Children later diagnosed with ADHD often show elevated activity, high negative emotionality, and early regulatory difficulties^23–26^. Anxiety and depressive disorders are linked to early behavioral inhibition and negative emotionality^27–30^. Increasing shyness across childhood has been associated with heightened risk for substance-use disorders^22^. Collectively, these findings suggest that variation in early temperament may signal vulnerability to specific mental health conditions^31,32^. However, prior work has largely focused on static trait levels rather than on how these temperament patterns change over time. Because temperament unfolds dynamically across childhood, single-time measurements may obscure meaningful differences in developmental trajectories. Departures from expected trajectories may thus capture patterns of change that are not reflected in static trait levels.

The availability of large-scale longitudinal cohorts and advances in statistical modeling now enable population-scale analyses of early-childhood temperament development, the identification of clinically relevant deviations from expected trajectories, and the characterization of age-dependent genetic influences. Here, we leverage data from the Norwegian Mother, Father and Child Cohort Study (MoBa), a prospective, genotyped, deeply phenotyped, population-based cohort^33,34^. We use mother-reported EAS measurements collected at 1.5, 3 and 5 years of age for more than 50,000 children, linked to nationwide health registry diagnoses extending from childhood into early adulthood. Using the FEMA-Long framework, a scalable mixed-effects approach designed for longitudinal population cohorts^35,36^, we model non-linear temperament trajectories across early childhood and distill each child’s trajectory into a single measure of persistent departure from developmental norms. Canonical Partial Least Squares (PLS) analysis then relates these deviations to a spectrum of mental and behavioral disorders. Furthermore, time-to-event modeling examines how early deviations are associated with the risk and timing of psychiatric disorders. Finally, by conducting genome-wide analyses of age-dependent genetic effects, we map how genetic influences on temperament change across development. Together, these analyses provide an integrated developmental framework that links early temperament trajectories and their genetic architecture to later mental health outcomes.

## Results

### Longitudinal structure of early-childhood temperament

We leveraged data from the MoBa Study, which prospectively follows more than 110,000 children with genetic profiling and comprehensive linkage to health registries. Our aim was to chart how early-childhood temperament trajectories relate to later mental health outcomes. In the present study, we used mother-reported temperament assessments collected when children were approximately 1.5, 3 and 5 years old (Fig. 1a). These assessments were based on the EAS instrument^14^, which quantifies four core temperament traits: *emotionality*, *activity*, *shyness*, and *sociability*. Temperament measurements from at least one time point were available for 50,529 children (45,028 at 1.5 years; 36,435 at 3 years; 25,969 at 5 years). These children were linked to data from the Norwegian Patient Registry (NPR), a nationwide registry of specialist-care diagnoses since 2008. Psychiatric diagnoses were defined according to ICD-10 Chapter V (Mental and behavioral disorders; F00–F99). Follow-up for psychiatric diagnoses extended through December 31, 2024. At the end of follow-up, participants were between 16 and 24 years of age, enabling capture of adolescent- and early adult-onset psychiatric diagnoses. Across the full sample of 50,529 children (Fig. 1b), the most prevalent condition was ADHD (F90.0; 3,211 cases), followed by moderate depressive episode (F32.1; 1,390 cases), nonorganic enuresis (F98.0; 1,361 cases), social phobia (F40.1; 1,165 cases) and Asperger syndrome (F84.5; 884 cases). In total, 372 unique ICD-10 codes including subcategories were observed in our sample (Supplementary Data 1).

**Figure 1:**
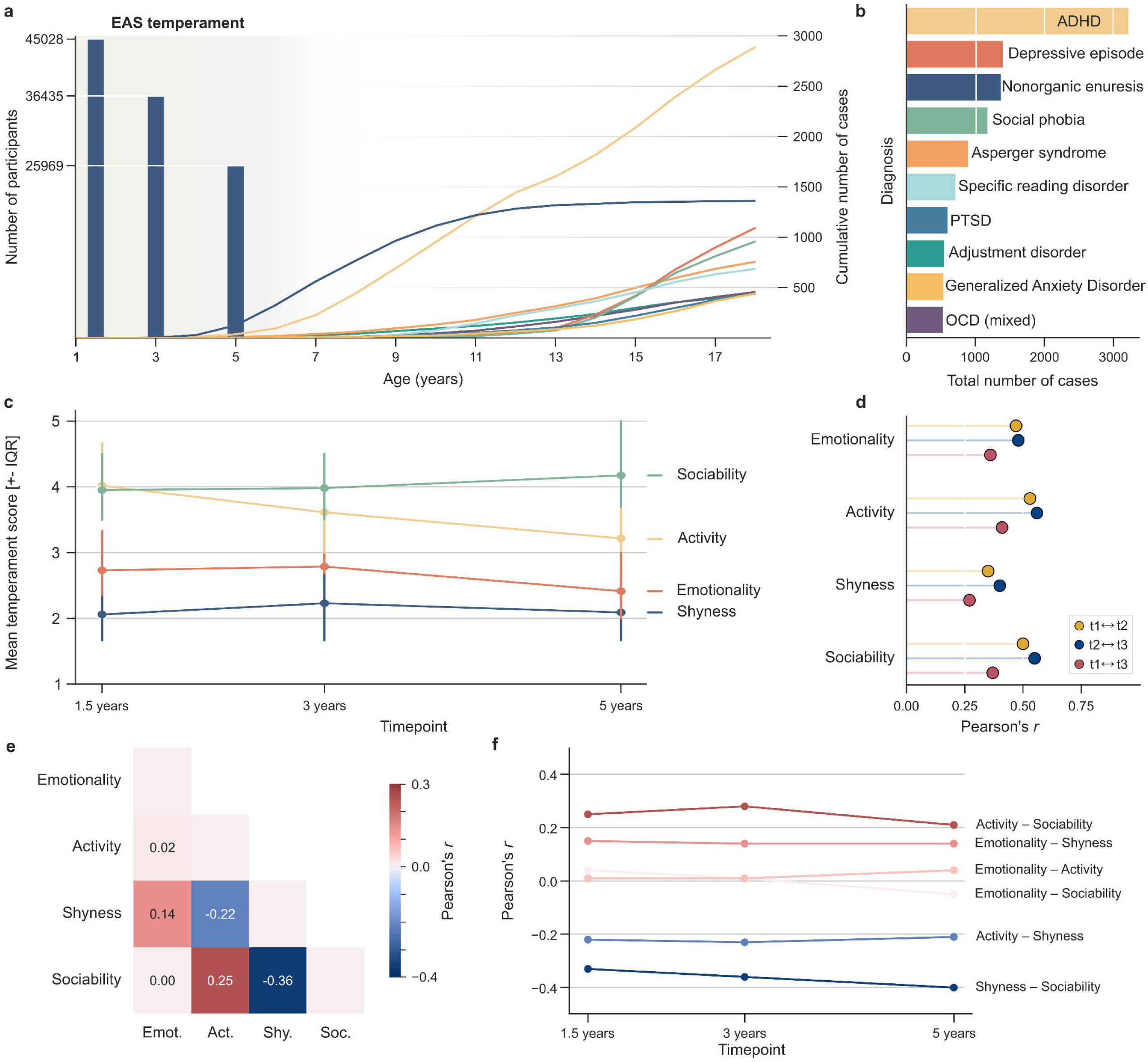
Temperament trajectories and diagnostic profiles in the MoBa subsample. **a.** Temperament measurements and diagnostic counts. Temperament (*emotionality*, *activity*, *shyness*, and *sociability*) data were available for 45,028 children at 1.5 years, 36,435 at 3 years and 25,969 at 5 years (left *y*-axis). The number of recorded diagnoses increased with age (right *y*-axis). Counts are cut off at age 18 years. Approximately 71% of the cohort had reached this age at the time of data extraction. **b.** Most common diagnoses. The ten most frequent mental and behavioral disorder diagnoses recorded until the end of 2024. Full ICD-10 codes are listed in Supplementary Data 1. ADHD corresponds to F90.0 (Disorders of activity and attention). PTSD refers to Post-traumatic stress disorder (F43.1), and OCD (mixed, F42.2) refers to Obsessive–compulsive disorder with mixed obsessions and compulsions. **c.** Temperament dynamics. Each temperament trait reflects the average of several 5-level questionnaire items. Points and error bars depict mean values and interquartile ranges at each timepoint. **d.** Temporal stability. Pearson correlations between all pairs of timepoints are shown for each temperament trait. Colors denote the three timepoint comparisons. **e.** Cross-trait similarity. Heatmap displays pairwise correlations between temperament traits, averaged across timepoints. **f.** Cross-trait similarity across time. Lines trace correlations between each pair of temperament traits. Line colors correspond to the mean correlation strength of each pair. Together, the four temperament traits show complementary dynamics that set the stage for examining their links to later diagnoses.

Each temperament trait was constructed as a composite score summarizing multiple questionnaire items with five response levels. All four traits showed moderate temporal stability across early childhood (Fig. 1c). Correlations ranged from *r* = 0.35 to 0.53 between ages 1.5 and 3 years, and from *r* = 0.40 to 0.55 between ages 3 and 5 years. *Activity* showed the highest stability and *shyness* the lowest across both intervals. Over the longer 1.5-to-5-year interval, correlations remained evident but weaker (*r* = 0.27–0.41; Fig. 1d). Temperament thus retains consistent individual differences while continuing to change.

In addition to temporal stability, we examined cross-trait relationships. The four temperament traits showed weak intercorrelations (Fig. 1e). Averaged across the three timepoints, the strongest relationship was observed between *shyness* and *sociability* (Pearson’s *r* = –0.36), whereas *sociability* and *emotionality* showed essentially no linear association (*r* = 0.00). These pairwise relationships were highly consistent across time, varying no more than *r* = 0.07 between any timepoint (Fig. 1f). Collectively, early-childhood temperament exhibited structured, rather stable patterns that form the foundation for the deviation analyses that follow.

### Deviations from population-based temperament trajectories

We leveraged the FEMA-Long framework^35,36^ to model age-dependent patterns in each temperament trait, accounting for repeated measurements and family structures inherent to population cohorts. The non-linear effect of age was modeled using natural cubic splines with unit height at knots (Fig. 2a; see Methods). In addition to the smooth effect of age, we included sex, genetic principal components and genotyping batch effects. Sex effects varied across traits, where *activity* showed the largest difference (Supplementary Data 2) since boys scored approximately 0.16 standard deviations higher than girls (β = –0.16 ± 0.01).

**Figure 2:**
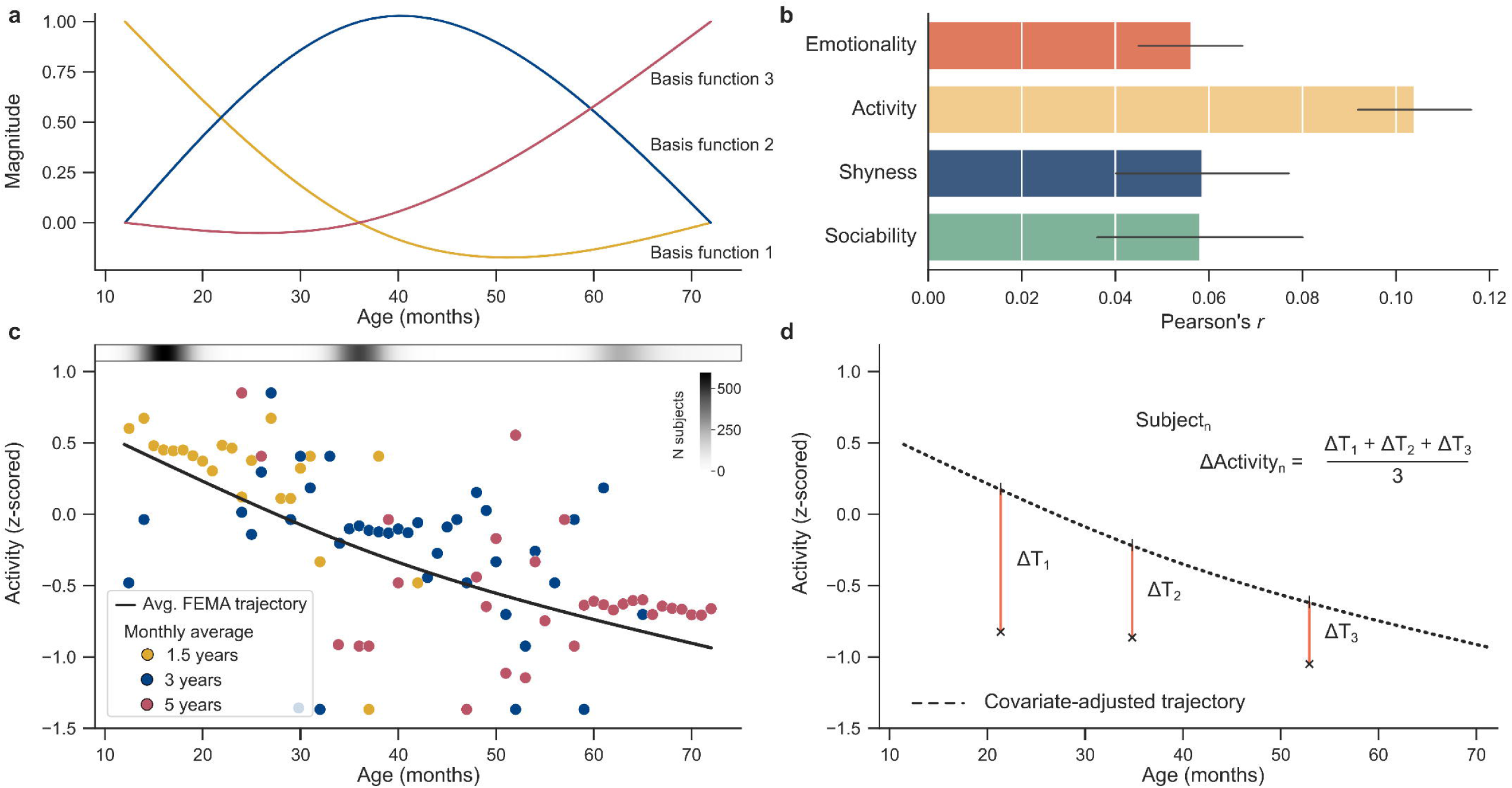
Deviations from predicted temperament trajectories. **a.** Spline basis functions modeling non-linear age effects. Age-dependent changes in temperament were modeled using natural splines with three basis functions. **b.** Model evaluation. Bar plots display correlations between predicted and observed temperament scores, averaged across timepoints. Error bars indicate the minimum and maximum correlations across timepoints. **c.** Modeled temperament trajectories. Mean *activity* scores are shown for each month with available data, colored by timepoint. The estimated population-level *activity* trajectory from the FEMA-Long model is overlaid. The grayscale bar indicates the number of observations available at each month. **d.** Deviations from trajectories. A schematic illustration of how temperament deviations were derived, based on the average difference between observed values and the covariate-adjusted predicted trajectory across the three timepoints. Collectively, the modeled trajectories capture normative temperament development while preserving substantial individual-level variation for quantification as deviations.

In the next step, we generated fitted values for each child using the estimated fixed effects. Fitted values represent the expected temperament scores based on age, sex, genetic principal components, and batch effect. As expected, fitted and observed scores values were only weakly correlated (*r* = 0.06–0.10 across timepoints; Fig. 2b), indicating that most variation in temperament lies beyond what is captured by the modeled covariates. Next, we defined temperament deviations as the difference between observed and fitted values (Fig. 2c, d). A positive deviation indicates that a child scores above expectation for their age and sex, a negative deviation the opposite. Deviations were then averaged across the three timepoints for the 18,651 children with complete data to emphasize consistent departures across early childhood rather than timepoint-specific fluctuations. This yielded four deviation measures that formed the basis for subsequent clinical analyses.

### Latent dimensions connecting temperament deviations and mental health

We examined whether deviations from population-level temperament trajectories in early childhood were linked to clinical diagnoses until early adulthood. To do so, we applied canonical PLS, a multivariate framework designed to extract latent dimensions that maximize shared variance between two sets of variables. In our case, PLS identified linear combinations of the four temperament deviations and linear combinations of all observed mental and behavioral disorders diagnoses (266 observed ICD-10 codes including subcategories) that covaried most strongly. This procedure yielded individual-level PLS scores that indicate how strongly each child expresses the multivariate pattern represented by each dimension.

Using permutation testing (Fig. S1), we identified two significant latent dimensions linking early-childhood temperament deviations to clinical diagnoses in early adulthood. To interpret each latent dimension, we examined the correlations between the original temperament deviations and the corresponding latent dimension scores (i.e., PLS scores). The first dimension reflected a temperament deviation profile marked by elevated *activity* (*r* = 0.71), and *emotionality* (*r* = 0.67), together with lower *sociability* (*r* = −0.38) (Fig. 3a). This dimension showed its strongest association with ADHD (*r* = 0.97), but was also associated with Asperger syndrome (*r* = 0.24), Tourette’s syndrome (*r* = 0.23), hyperkinetic conduct disorder (*r* = 0.23), and specific reading disorder (*r* = 0.22).

**Figure 3:**
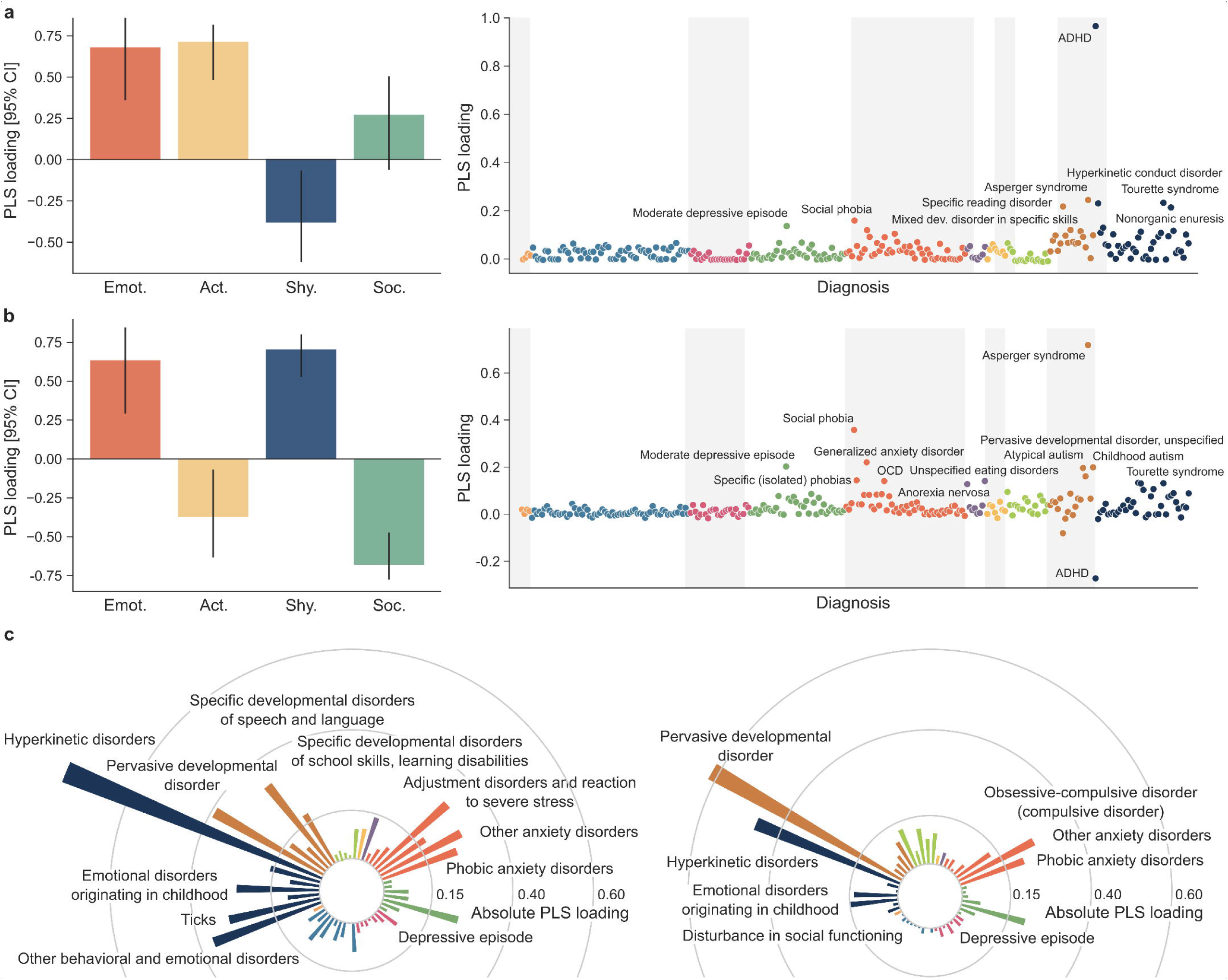
Multivariate links between temperament deviations and diagnoses. Canonical Partial Least Squares (PLS) analysis identified latent dimensions capturing shared variation between temperament deviations and mental and behavioral disorders diagnoses. Temperament loadings were defined as correlations between temperament PLS scores and the four deviation measures, and diagnosis loadings as correlations between diagnosis PLS scores and 266 ICD-10 subcategory diagnoses. **a**. First dimension spotlights ADHD. Temperament loadings for the first latent temperament dimension are shown with 95% confidence intervals obtained from 10,000 bootstrap resamplings (left). Diagnosis loadings are displayed on the right, grouped and colored according to ten overarching diagnostic categories. **b**. Second dimension highlights Asperger syndrome and social phobia. Temperament and diagnosis loadings for the second latent temperament dimension are shown using the same conventions as above. **c.** Hyperkinetic disorders dominate diagnosis association. Diagnosis loadings are shown for models fitted to ICD-10 diagnostic categories, for the first dimension (left) and second dimension (right). Together, two latent dimensions link distinct patterns of temperament deviations with structured diagnostic profiles across childhood and adolescence.

The second dimension reflected a distinct temperament deviation profile characterized by elevated *shyness* (*r* = 0.70) and *emotionality* (*r* = 0.63), together with reduced *sociability* (*r* = −0.67) and *activity* (*r* = −0.37) (Fig. 3b). This dimension was associated with increased risk for Asperger syndrome (*r* = 0.72), social phobia (*r* = 0.36), generalized anxiety disorder (*r* = 0.22), and major depressive disorder (*r* = 0.31). Notably, it showed a negative relationship with ADHD (*r* = −0.27). Taken together, the two latent dimensions captured orthogonal, data-driven patterns of temperament deviation from the developmental norm during childhood, each differently associated with psychiatric outcomes in adolescence and early adulthood.

To assess the robustness of these associations, we repeated the analysis using ICD-10 category codes, which aggregate the 266 subcategory codes into 50 broader diagnostic categories (Fig. 3c). Rerunning the PLS model yielded again two significant dimensions with temperament profiles similar to those obtained in the full analysis (Fig. S2). The first latent dimension was dominated by hyperkinetic disorders (*r* = 0.86), the major-category umbrella for all ADHD subtypes, followed by pervasive developmental disorder (*r* = 0.39) and other behavioral and emotional disorders that usually occur in childhood and adolescence (*r* = 0.36). The second dimension showed the highest loadings for pervasive developmental disorder (*r* = 0.68), hyperkinetic disorders (*r* = −0.48) and other anxiety disorders (*r* = 0.26). As a sensitivity analysis, we repeated this analysis using diagnoses from both the NPR (specialist healthcare) and the KUHR database (Norwegian Control and Payment of Health Reimbursements Database, see Methods). KUHR captures reimbursed healthcare encounters including primary care consultations. Results were consistent with the NPR-only analysis, as very few additional diagnoses were present in KUHR alone (Fig. S3). All subsequent analyses therefore used NPR diagnoses only. All associations and confidence intervals (CIs) are summarized in Supplementary Data 3. Collectively, these results reinforced the strong links between deviations from temperament trajectories and mental and behavioral diagnoses.

### Temperament deviations and the timing of ADHD diagnosis

Having identified two diagnostically relevant latent dimensions, we examined their association with the risk and timing of first recorded diagnoses. ADHD was selected for follow-up time-to-event analyses because it showed the most consistent and robust associations across both latent dimensions in the multivariate model (Fig. S4). To quantify this association, we modeled time to first ADHD diagnosis using Cox proportional hazards models with age in months as the underlying time scale. The two latent dimension scores were included as predictors together with relevant covariates (Table S1), allowing us to estimate how variation along each latent dimension related to the hazard of receiving an ADHD diagnosis.

Only the first latent dimension showed a significant association with increased hazard of ADHD diagnosis (PLS 1: hazard ratio (HR) = 1.52, 95% CI 1.41–1.65), whereas the second dimension was not significant (PLS 2: HR = 1.05, 95% CI 0.98–1.12; Fig. 4a). Kaplan–Meier survival curves showed a clear increase in cumulative risk with high scores on the first dimension (Fig. 4b). Cumulative risk also differed by sex, with males showing consistently higher risk and the sex difference widening from early childhood onward (Fig. 4c).

**Figure 4:**
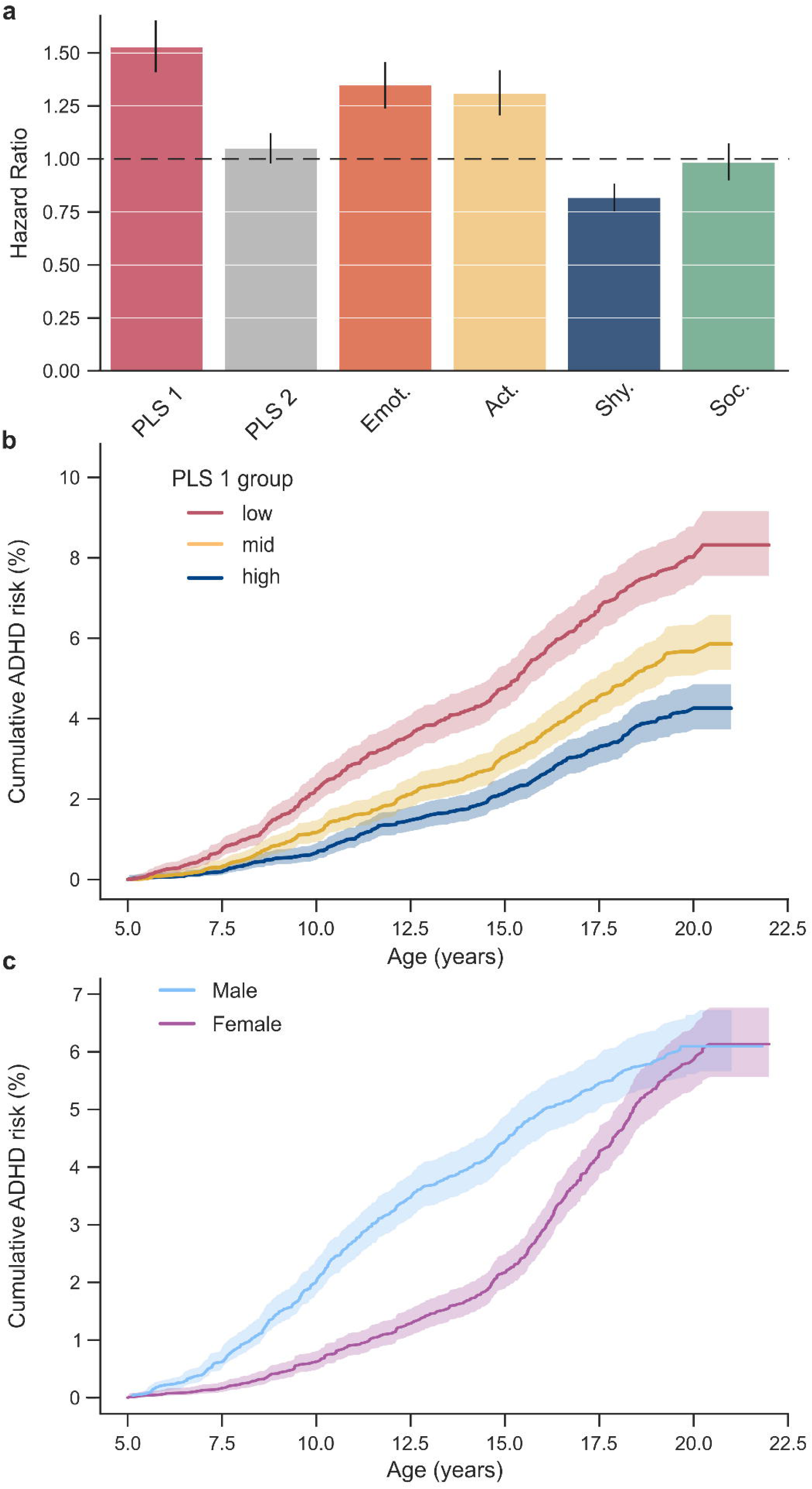
Decoding ADHD diagnosis using latent dimensions. **a.** ADHD risk across temperament traits. Hazard ratio of the latent temperament dimensions is displayed along with hazard ratios of temperament deviations. Error bars depict 95% confidence interval. **b.** ADHD risk and first latent temperament dimension. Kaplan–Meier curves show cumulative ADHD risk for three groups stratified by the first latent dimension. Curves correspond to low, intermediate, and high scores on the first dimension. Cumulative risk is calculated as one minus the probability of remaining ADHD-free up to each age. **c.** Sex-specific ADHD risk. Kaplan–Meier curves display the cumulative probability of receiving an ADHD diagnosis separately for males and females, reflecting sex-specific differences in ADHD risk across development. Taken together, latent dimensions capture variation in ADHD risk and are associated with differences in the timing of diagnosis.

Although the temperament deviation dimensions were not optimized specifically for ADHD diagnosis, the hazard ratio associated with the first latent dimension score appeared to exceed those observed for individual temperament deviations (Fig. 4a). The higher hazard ratio indicates that the latent dimension efficiently captures the multivariate temperament signal relevant to ADHD risk. Given the prominence of ADHD in both latent dimensions, time-to-event analyses initially focused on this diagnosis. For comparison, we applied the same time-to-event analysis pipeline to Asperger syndrome and social phobia, diagnoses that were prominent in the second latent dimension. Both latent dimensions were associated with increased hazard of Asperger syndrome (PLS 1: HR = 1.16, 95% CI 1.01–1.34; PLS 2: HR = 1.60, 95% CI 1.42–1.82). In contrast, only the second latent dimension was associated with increased hazard of social phobia (HR = 1.19, 95% CI 1.07–1.32; Fig. S5). Notably, females exhibited markedly higher cumulative risk of social phobia across childhood and adolescence compared to males (Fig. S5). Together, these findings showed that deviations from temperament trajectories are associated with both the risk and timing of psychiatric diagnosis.

### Genetics of temperament trajectories

The temperament deviations examined above reflect individual differences in how children depart from population-level developmental norms. Because these deviations capture variation in the underlying temperament traits, we next examined the genetic architecture of temperament itself. The FEMA-Long framework decomposes the total phenotypic variance into individual-level, family-level and additive genetic variance components, each of which can vary across traits and over time within a trait (Fig. S6). Focusing on the genetic component revealed variation in its relative contribution across the four traits (Fig. 5a). For example, the proportion of variance in *activity* attributable to the genetic component increased from 0.41 at 1.5 years, to 0.50 at 3 years, and then decreased to 0.46 at 5 years. Overall, these estimates indicate that the relative contribution of additive genetic variance remains largely stable across early childhood, with only modest fluctuations over time.

**Figure 5:**
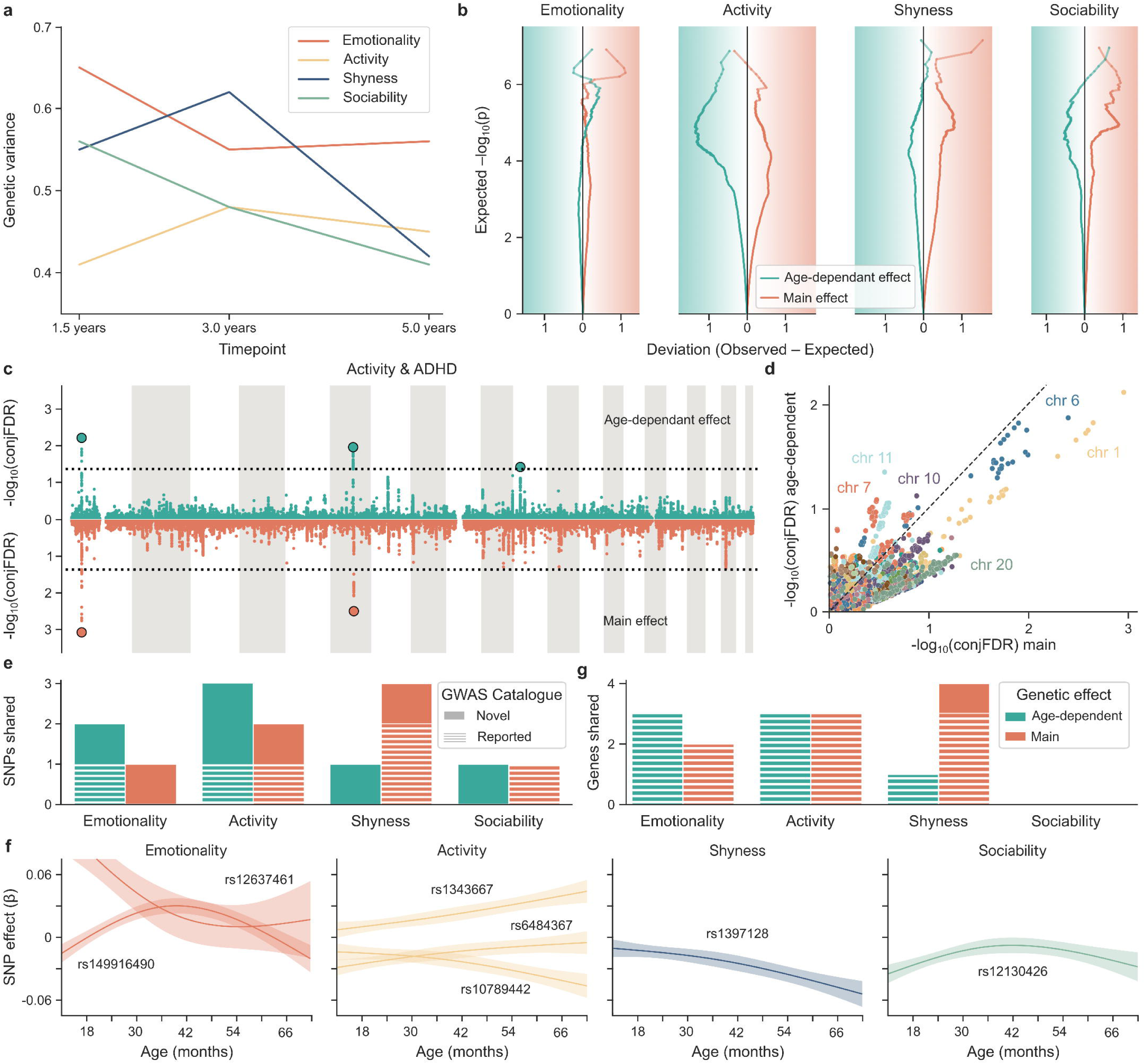
Dynamic genetic effects linking early temperament and ADHD. **a.** Temporal dynamics of genetic variance. Genetic variance estimates for each temperament trait at 1.5, 3, and 5 years, as derived from FEMA-Long. **b.** Enrichment of GWAS signal across main and age-dependent genetic effects. Each panel shows a deviation-based Q-Q plot, where the *x*-axis represents the difference between observed and expected –log_10_(*p*). Curves show main genetic effects and age-dependent effects (see legend). Leftward deviation of the main-effect curve indicates enrichment of main genetic effects, whereas rightward deviation of the longitudinal-effect curve indicates enrichment of age-dependent effects. Deviations in the opposite direction reflect deflation. **c.** Shared SNPs between *activity* and ADHD. Miami plots show SNPs jointly associated with ADHD and *activity* for the main (bottom) and age-dependent (up) genetic effects. The *y*-axis displays −log_10_ conjFDR *p*-values. Black circles mark significant lead SNPs within linkage disequilibrium blocks. **d.** Comparison of association strength for main and age-dependent effects. Transformed conjFDR *p*-values for main and age-dependent genetic effects are plotted for all SNPs. **e.** Lead SNP overlap between temperament traits and ADHD. Bar plots show the number of significant lead SNPs jointly associated with ADHD and each temperament trait, separately for main and age-dependent effects. Dashed segments indicate SNPs previously reported in the GWAS Catalog. **f.** Trajectories of age-dependent effects. Estimated age-dependent genetic effects are shown across age for all significant lead SNPs identified in the conjFDR analyses. **g.** Gene-level overlap between temperament traits and ADHD. Bar plots show the number of genes corresponding to lead SNPs shared between ADHD and each temperament trait. Together, the results highlight dynamic genetic influences on temperament, with several effects emerging from genomic regions distinct from those implicated by age-independent (main) genetic effects.

To investigate genetic influences on temperament trajectories, we applied the FEMA-Long framework to perform genome-wide association analyses (GWAS) that estimate both main and age-dependent single nucleotide polymorphism (SNP) effects. Unlike standard cross-sectional GWAS, this approach allows genetic effects to vary non-linearly across development. For each temperament trait, we modeled every SNP with both a main effect and an interaction with smooth age basis functions (see Methods). Each GWAS thus quantified the overall genetic effect of each SNP and whether that effect changed across early childhood. The resulting Q-Q-plots indicated polygenic enrichment (Fig. 5b, Fig. S7), though few SNPs reached genome-wide significance (*p* < 5×10□□, Fig. S8).

The strong phenotypic association between early temperament and ADHD suggested potential shared genetic architecture. To investigate this possibility, we applied conjunctional false discovery rate (conjFDR) analysis^37,38^ to identify SNPs jointly associated with ADHD and each temperament trait, considering both main genetic effects and age-dependent effects (Figs. S9–S10; Table S2). As an illustrative example, three loci with significant main effects and two loci with significant age-dependent effects were shared between *activity* and ADHD (Fig. 5c). Comparing the conjFDR significance of main and age-dependent effects revealed both shared and distinct patterns of genetic association (Fig. 5d). Specifically, lead SNP rs6484367 on chromosome 11 showed predominantly age-dependent effects, whereas rs10789442 on chromosome 1 displayed a stronger main effect. Lead SNP rs1343667 on chromosome 6 showed evidence of both components. These findings highlight the contribution of age-dependent effects to the shared genetic architecture between temperament and ADHD.

Across the four temperament traits, *activity* showed the highest number of SNPs shared with ADHD (Fig. 5e). For lead SNPs with significant age-dependent effects in the conjFDR analyses, we reconstructed their effect trajectories across early childhood using the spline-based interaction terms from the fitted FEMA-Long model (Fig. 5f). Several SNPs, such as rs1368742 (*emotionality*) and rs6484367 (*shyness*), displayed decreasing effect sizes across age. In contrast, other SNPs (e.g., rs1343667 for *activity* and rs1397128 for *shyness*) showed effect sizes that increased with age. Notably, these SNPs have no previously reported associations in the GWAS Catalog^39^. Another SNP with an increasing negative effect size (rs10789442 for *activity*) was previously associated with self-reported mathematical ability.

Finally, we mapped lead SNPs from the conjFDR analysis to genes based on genomic position (FUMA; see Methods) and identified eight genes across all temperament traits (Fig. 5g, Table S2). *ST3GAL3* was implicated across four trait–effect combinations (*activity* main and age-dependent effects, *shyness* main effect, *emotionality* main effect), suggesting a broad contribution to early temperament variation. Two additional genes, *KDM4A* and *PTPRF*, were each implicated across three combinations (*activity* main and age-dependent effects, *shyness* main effect). The remaining genes (*CADM2*, *CDH8*, *SLC6A9*, *ZNF462*, and *RP11-508N12.4*) were each associated with two or fewer trait–effect combinations. GWAS Catalog queries indicated that seven of the eight genes have previously been associated with cognitive, educational, psychiatric, or neurodevelopmental phenotypes, including educational attainment, socioeconomic status, ADHD, autism spectrum disorder, and intelligence (Fig. S11). *RP11-508N12.4*, which was not represented in the GWAS Catalog, has been linked to migraine in a recent large-scale study^40^. Collectively, these results identify loci not previously reported in the GWAS Catalog and characterize how genetic effects at both novel and previously reported loci vary across early childhood.

## Discussion

In this study, we leveraged a longitudinal birth cohort to characterize how early-childhood temperament deviations relate to psychiatric outcomes in early adulthood. We identified two complementary temperament deviation dimensions associated with risk profiles spanning multiple diagnoses, including ADHD, Asperger syndrome, and social phobia. Notably, ADHD was linked to both dimensions, which suggests that the disorder may arise from multiple developmental pathways. Additionally, we uncovered a shared genetic basis between temperament trajectories and ADHD. By modeling temperament trajectories, we identified age-dependent genetic effects at shared loci. Genetic liability for psychopathology thus appears to be developmentally regulated, with effects that may be missed by cross-sectional approaches.

Temperament is only moderately stable across early childhood^41^ (*r* = 0.27–0.53 in our data), meaning that a single measurement cannot distinguish a child’s stable behavioral tendency from age-specific fluctuation. By modeling temperament across three assessments, we evaluated each child against population-level developmental norms and averaged the resulting deviations across timepoints. These deviations quantify what clinicians observe informally: some children are consistently more active or emotional than expected for their age and sex. Whereas prior approaches linked individual traits to specific disorders at single timepoints^10^, entering deviations into a multivariate analysis revealed that coordinated patterns across traits mapped onto distinct patterns of psychiatric risk. More pronounced deviations were associated with a higher hazard of diagnosis. The trajectory-deviation approach thus captures dimensions of transdiagnostic risk that are invisible to single-timepoint, single-trait designs.

The two identified temperament deviation dimensions captured distinct but complementary clinical patterns. In the first, elevated *activity* and *emotionality* alongside reduced *sociability* were associated with ADHD and related neurodevelopmental diagnoses including Tourette’s syndrome, hyperkinetic conduct disorder, and specific reading disorder. These co-occurring diagnoses are consistent with the high comorbidity observed between ADHD and other neurodevelopmental conditions^42,43^. In the second, elevated *shyness* and *emotionality* alongside reduced *activity* and *sociability* were most strongly associated with Asperger syndrome, social phobia, and generalized anxiety, capturing a shared phenotype of social withdrawal that spans neurodevelopmental and internalizing boundaries^30,44,45^. What differentiated the two profiles was *activity* versus *shyness*: elevated *activity* characterized the externalizing-like profile, while elevated *shyness* characterized the social withdrawal profile. *Emotionality* was elevated in both, which suggests a transdiagnostic role that cuts across domains^46–48^. These complementary patterns align with hierarchical models that frame early developmental variation as broad vulnerability rather than disorder-specific risk^49^.

ADHD offers a clear illustration of this broad vulnerability pattern. It was linked to both latent dimensions, but with elevated *emotionality* in the first and reduced *emotionality* in the second. This means that children arriving at the same ADHD diagnosis depart from expected temperament development in opposite directions on *emotionality*: some through heightened emotional reactivity, others through reduced reactivity. This divergence may help explain the disorder’s well-documented clinical heterogeneity^42,43^. If distinct temperament profiles converge on the same diagnosis, then apparent heterogeneity within ADHD may partly reflect different developmental origins. These origins may later diverge into different comorbidity patterns, which may help explain why ADHD so frequently precedes more differentiated psychiatric outcomes^50^. The strength of the phenotypic link between temperament deviations and ADHD raises the question of whether they also share genetic architecture.

Using the FEMA-Long framework to estimate age-dependent genetic effects, we identified genetic loci jointly associated with temperament trajectories and ADHD. These variants exhibited both main and age-dependent effects on temperament across early childhood. The presence of both types of effects is consistent with classical quantitative genetic evidence that genetic factors contribute to both stability and change in temperament during development^17,51^. It also aligns with longstanding distinctions between genetic influences that support continuity and those associated with developmental change^3,52,53^. Our findings extend this work at the molecular level. Shared genetic influences between temperament and ADHD are not uniform. Some loci showed predominantly main effects, others predominantly age-dependent effects, and some showed evidence of both. Moreover, the direction of age-dependent effects varied, with some increasing and others decreasing. Together, these results indicate that the shared genetic architecture between temperament and ADHD is partly dynamic, with effects that would be missed by analyses assuming static genetic effects.

The implicated genes have been previously associated with cognitive ability, educational attainment, and multiple psychiatric conditions beyond ADHD. This suggests that the genetic overlap between temperament and ADHD is embedded within a broader architecture spanning cognition, self-regulation, and neurodevelopmental risk^54,55^. The genetic breadth parallels the phenotypic picture, where different temperament profiles converge on the same diagnostic outcomes. Furthermore, the wide phenotypic reach of the identified loci suggests that age-dependent genetic effects extend beyond the temperament-ADHD link alone. More broadly, genetic liability for ADHD, and likely other disorders, appears to be developmentally regulated.

A key question is whether current approaches to genetic risk prediction adequately capture such developmental dynamics. Polygenic risk scores represent the primary tool for translating genetic findings into individual risk prediction. They aggregate the effects of many variants into a single measure of liability^56^. Current implementations, however, typically assume that these genetic effects are constant across development^57^. As such, they may not capture the developmentally varying influences identified here. Incorporating age-dependent genetic effects into risk prediction frameworks could distinguish children whose genetic risk is expressed early versus later. This would in turn enable more precisely timed monitoring or intervention.

For clinical practice, monitoring deviations from expected developmental trajectories may offer an additional signal for identifying children at elevated risk before formal diagnosis. Much like pediatricians track height and weight against population norms and flag children who deviate, repeated temperament assessment could allow clinicians to plot a child’s development against age-expected trajectories, with persistent departures signaling the need for follow-up. In our analyses, such deviations were associated with an increased hazard of psychiatric diagnosis across childhood and adolescence. Risk patterns differed by sex, with males showing higher cumulative ADHD risk and females a higher risk for social phobia. These patterns mirror well-established sex differences in the prevalence of both disorders^58,59^ and underscore the importance of incorporating biological and environmental context into monitoring frameworks^60^. Temperament questionnaires such as the EAS are brief and inexpensive. Their integration into routine developmental monitoring is therefore feasible in principle. Prospective studies are now needed to test whether deviation-based monitoring, combined with age-dependent polygenic risk scores, can translate into earlier identification and improved outcomes.

Some limitations should be considered when interpreting our findings. Registry-based psychiatric diagnoses capture clinically recognized disorders but lack symptom-level granularity and reflect administrative timing rather than biological onset, which may introduce lag into survival estimates. The birth cohort design favors capture of early-onset diagnoses like ADHD over later-onset conditions like bipolar disorder or schizophrenia, and cross-diagnostic comparisons are therefore limited. Temperament was assessed through parent reports, which may be influenced by reporting biases. Our longitudinal, deviation-based approach partially mitigates this concern by emphasizing within-individual change rather than absolute levels, although systematic reporting biases could still influence trajectory estimates. Loss to follow-up in this cohort is non-random and can lead to both attenuation and inflation of associations^61^. Additionally, temperament measures may overlap with early symptoms of the disorders they predict. *Activity* items, for instance, resemble early ADHD symptoms. However, this reflects genuine continuity between normal temperament variation and clinical disorder^62^ rather than a methodological artifact. Finally, the cohort is drawn from a European population-based sample and genetic analyses were restricted to participants of European ancestry. Findings may therefore have reduced generalizability to populations with different ancestral backgrounds, healthcare systems, or sociocultural contexts.

In conclusion, we show that deviations from expected temperament trajectories in early childhood are associated with psychiatric diagnoses across neurodevelopmental and internalizing domains. ADHD emerged as a particularly salient outcome, associated with distinct patterns of early temperament deviation. These associations are underpinned by shared genetic architecture between temperament and ADHD that is itself partly dynamic, with effects that vary across early childhood. Together, these findings suggest that early psychopathology risk is best understood not as the presence of specific traits, but as deviations from expected trajectories shaped by dynamically expressed genetic influences. Linking trajectory deviations to long-term psychiatric risk supports a shift from reactive diagnosis toward proactive, developmentally informed monitoring of mental health risk.

## Methods

### MoBa: A nationwide population cohort

The Norwegian Mother, Father and Child Cohort Study (MoBa) is a population-based pregnancy cohort study conducted by the Norwegian Institute of Public Health^33^. Participants were recruited from all over Norway from 1999–2008. The women consented to participation in 41% of the pregnancies. The cohort includes approximately 114,500 children, 95,200 mothers and 75,200 fathers. All participants provided written informed consent; children are included after consent from the mother, once they reach 18 years old. MoBa requires informed consent from the participant for further use and storage of the data. Umbilical cord blood collected at birth was used for genotyping^63^. MoBa was linked to the Medical Birth Registry of Norway (MBRN), a compulsory national registry containing clinician-reported information on all Norwegian births. The present study used version 12 of the quality-assured maternally reported questionnaire data, released in January 2019. Linkage to the Norwegian Patient Registry (NPR) provided ICD-10 diagnoses from 2008 through 2024, enabling longitudinal follow-up of health outcomes. Analyses were restricted to participants who had not withdrawn consent from MoBa as of April 25, 2025. Quality-controlled genotype data for 75,807 children were obtained from the MoBaPsychGen pipeline v115 (described previously^34^). Sex assigned at birth and birth year were extracted from the 6-month questionnaire. Family membership was defined by maternal identifier, such that children sharing the same mother were treated as a single family unit.

The establishment and data collection of MoBa were approved by the Norwegian Data Protection Agency and the Regional Committees for Medical and Health Research Ethics (REK). MoBa operates under the Norwegian Health Registry Act. The current study was approved by the MoBa administrative board at the Norwegian Institute of Public Health (NIPH) and REK (reference 2016/1226/REK Sør-Øst C).

### Temperament assessments across early childhood

Temperament was assessed using maternally reported Emotionality, Activity, Shyness, and Sociability Temperament questionnaire (EAS)^13,14^ administered when children were approximately 1.5, 3 and 5 years old. The EAS measures four temperament traits: *emotionality* (irritability/anger), *activity* (activity level), *shyness* (fear), and *sociability* (positive affect/including approach). The original 20-item questionnaire assesses these four traits using five items per trait. In MoBa, only 11 items were administered consistently across all three timepoints. Analyses were therefore restricted to these items (listed in Table 1). Mothers rated each item on a five-point scale from “not at all typical” to “very typical” of their child. For each temperament trait, we computed summary scores by averaging available item responses. Analyses included children with available genotype data, recorded biological sex, and at least one valid EAS assessment. Valid assessments were defined as questionnaires completed when the child was aged 1–6 years with no missing items. The sequential application of these criteria and resulting sample sizes are detailed in Table 2. After all exclusions, 45,028 children contributed temperament data at 1.5 years, 36,435 at 3 years, and 25,969 at 5 years, with 18,651 children measured at all three timepoints.

**Table 1:**
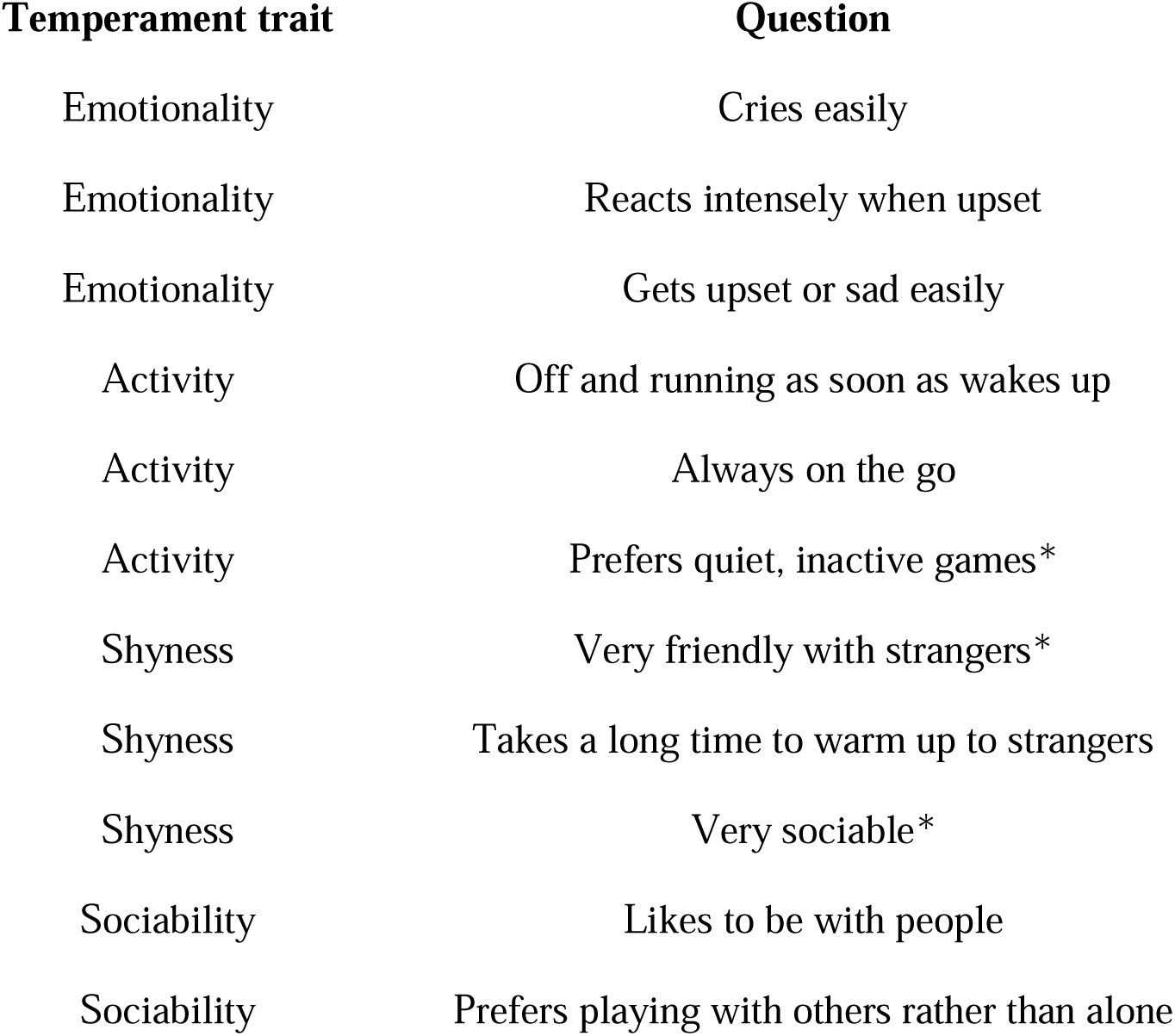
EAS questionnaire items and corresponding temperament traits. Each item is mapped to one of the EAS domains (*emotionality*, *activity*, *shyness*, *sociability*). Items marked with an asterisk were reverse-scored so that higher values consistently indicate higher levels of the corresponding temperament trait. All items were rated on a 1–5 scale.

| Temperament trait | Question |
| --- | --- |
| Emotionality | Cries easily |
| Emotionality | Reacts intensely when upset |
| Emotionality | Gets upset or sad easily |
| Activity | Off and running as soon as wakes up |
| Activity | Always on the go |
| Activity | Prefers quiet, inactive games* |
| Shyness | Very friendly with strangers* |
| Shyness | Takes a long time to warm up to strangers |
| Shyness | Very sociable* |
| Sociability | Likes to be with people |
| Sociability | Prefers playing with others rather than alone |

**Table 2:**
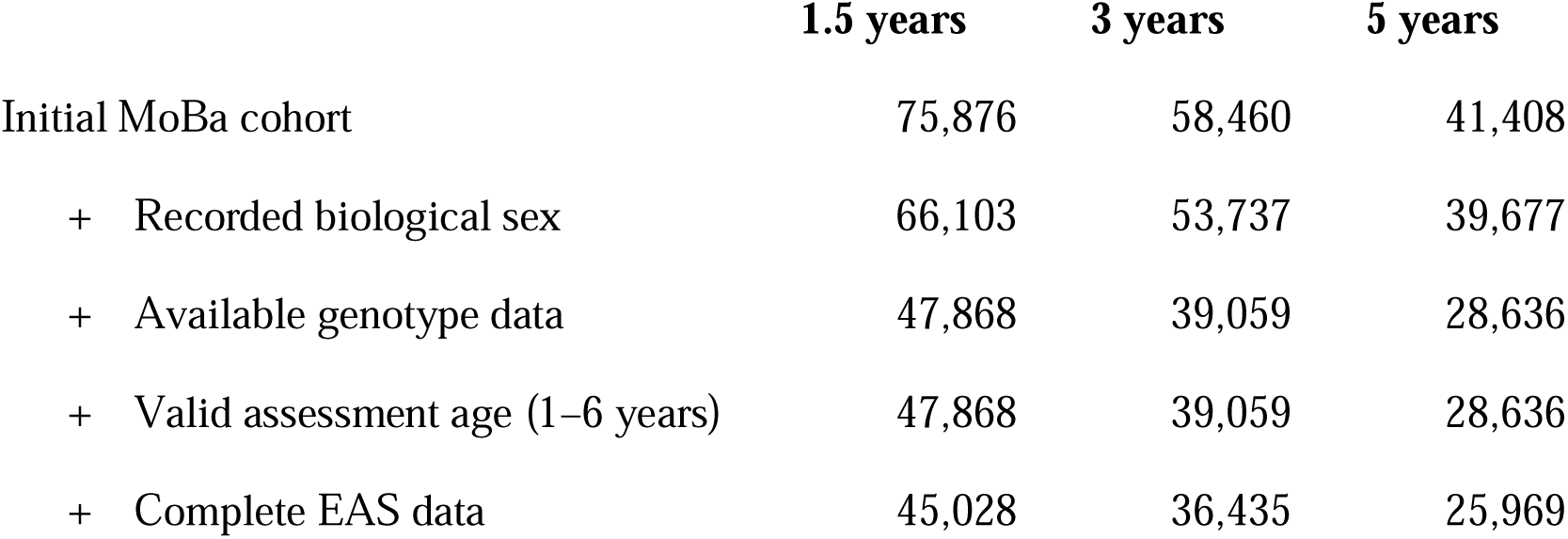
Sample sizes after application of inclusion criteria. Sample sizes at each temperament assessment wave (1.5, 3, and 5 years) after applying each inclusion criterion sequentially. Criteria included: available genotype data from MoBaPsychGen pipeline v115^34^, recorded biological sex, valid assessment age (child aged 1–6 years at questionnaire completion), and complete EAS questionnaire data (all items answered). The final row shows the analysis sample contributing valid temperament data at each timepoint. Of the final analysis sample, 18,651 children contributed valid temperament data at all three timepoints.

|  | 1.5 years | 3 years | 5 years |
| --- | --- | --- | --- |
| Initial MoBa cohort | 75,876 | 58,460 | 41,408 |
| + Recorded biological sex | 66,103 | 53,737 | 39,677 |
| + Available genotype data | 47,868 | 39,059 | 28,636 |
| + Valid assessment age (1–6 years) | 47,868 | 39,059 | 28,636 |
| + Complete EAS data | 45,028 | 36,435 | 25,969 |

### Diagnostic data from the Norwegian Patient Registry

Diagnoses were obtained from the NPR^64^, which records ICD-10 diagnostic codes^65^ from specialist healthcare encounters in Norway, including inpatient and outpatient hospital services and privately contracted specialist care. In this study, we focused on mental and behavioral disorders (ICD-10 codes, chapter V: F00–F99), as access to somatic diagnoses was limited. Diagnostic information was available from January 2008 through December 2024. The NPR linkage procedure used in the present MoBa analyses is described in detail in^66^. A complete list of diagnoses and their frequencies is provided in Supplementary Data 1. We also extracted diagnoses from the KUHR database (Norwegian Control and Payment of Health Reimbursements Database), which contains reimbursement records for healthcare encounters funded through the Norwegian National Insurance Scheme, including primary care consultations and some specialist services not captured by NPR. Notably, these nationwide records are available from 2006, compared with 2008 for NPR. Yet, KUHR contributed only a few additional diagnoses beyond those already recorded in NPR (Fig. S3). Given this observation, and because NPR diagnoses reflect specialist healthcare assessments with more comprehensive clinical evaluation, all main analyses were restricted to NPR diagnoses.

ICD-10 codes follow a hierarchical structure ranging from broad categories to increasingly specific subcodes. We used both detailed ICD-10 codes (four- and five-character subcategory codes, e.g., F90.1) and the corresponding higher-level code categories (e.g., F90), depending on the analytic context. For each condition, the date of first recorded diagnosis was used in all time-to-event analyses.

### Longitudinal modeling of temperament trajectories

Temperament trajectories were modeled using the FEMA-Long framework v3.0.0^35,36^. FEMA-Long is a scalable mixed-effects modeling framework specifically designed for longitudinal population cohorts with correlated observations. The method extends classical mixed-effects modeling by allowing random effects to vary over time, which enables flexible estimation of the covariance structure without imposing restrictive parametric assumptions on temporal dependence. In the present study, random effects were specified to capture individual, family, and genetic similarity, enabling decomposition of variance into individual, familial, and genetic components.

Each temperament trait was modeled separately. The covariates included age (modeled nonlinearly as described below), biological sex, the first twenty genetic principal components^34^, and genetic batch indicators. All continuous covariates were mean-centered and scaled to unit variance prior to modeling. Covariate effects, except for age, were modeled as time-invariant fixed effects.

Rather than specifying age as a strictly linear effect, we used a natural cubic spline basis expansion of age with an interior knot at 3 years and boundary knots at the minimum and maximum observed ages, ensuring adequate flexibility across the early-childhood window. To avoid collinearity with the intercept, the three basis functions were reparameterized via singular value decomposition (SVD), yielding two orthogonal components that, together with the intercept, span the same function space and ensure a full-rank design matrix.

Genome-wide genetic relatedness was captured via a genetic relationship matrix computed across 500,243 SNPs that were either directly genotyped in at least one imputation batch or had an imputation INFO score exceeding 0.99785 across all imputation batches^34^. This matrix was included as a random-effect covariance component within the FEMA-Long framework.

Genome-wide association analyses were performed using FEMA-Long framework applied to 6,981,748 SNPs^34^. For each temperament trait, FEMA-Long estimated the main effect of the SNP, as well as the age-dependent SNP effect (modeled as an interaction of the SNP with the smooth basis functions). To test whether a SNP exhibited a significant age-varying effect, we performed an omnibus Wald test jointly testing all spline coefficients associated with that SNP (contrast matrix equal to the identity over the spline terms). Manhattan plots display *p*-values for the main effect and Wald-test *p*-values for the age-dependent effect, allowing visual comparison of static and dynamic genetic influences on temperament. Genome-wide significance was defined using the conventional threshold of 5×10□□.

### Multivariate analysis of temperament-diagnosis associations

#### Constructing the inputs for multivariate analysis

We quantified temperament deviations for the 18,651 children with complete measurements across all three timepoints (Fig. S12). For each child, we generated expected temperament values using the fixed-effect component of the FEMA-Long model, which estimates fixed-effects coefficients while accounting for the covariance structure induced by the random effects. Deviations were then computed as the signed difference between observed and model-estimated temperament values and averaged across the three timepoints. We retained the signed differences to capture persistent upward or downward deviations from the expected developmental trajectory. This yielded a single deviation score capturing the extent to which a child consistently differed from the model-estimated developmental trajectory. Resulting deviation measures served as the basis for linking temperament dynamics to healthcare diagnoses.

To obtain a stable, low-dimensional representation of the diagnostic data, we applied principal component analysis (PCA) to the binary ICD-10 matrix. We retained components accounting for 95% of the variance, which corresponded to the first 100 components (Fig. S1). This resulted in two aligned input matrices for subsequent multivariate modeling: an 18,651 × 4 matrix of temperament deviations and an 18,651 × 100 matrix of diagnostic components.

### Latent dimensions linking temperament and diagnosis

We leveraged canonical Partial Least Squares (PLS) to identify multivariate associations between temperament deviations and diagnostic profiles^67^. PLS extracts latent dimensions that maximize the covariance between the two high-dimensional datasets. These dimensions are orthogonal and ordered by the amount of cross-covariance they explain, with the first dimension capturing the dominant shared structure.

To determine how many PLS dimensions reflected signal rather than noise, we compared the observed covariance of each dimension with a permutation-derived null distribution. We generated the null distribution by repeating the full PLS procedure 10,000 times. In each iteration, we permuted the temperament matrix at the family level by shuffling family blocks (keeping siblings together), while leaving the diagnostic data unchanged, thereby preserving within-family dependence. For each iteration, we recorded the covariance explained by the first PLS dimension, yielding a null distribution of covariances expected under no association. For each observed PLS dimension, the *p*-value was computed as the proportion of permutations in which the permuted covariance exceeded the observed covariance. Dimensions with *p* < 0.05 were retained (Fig. S1).

### Phenotypic contributions to latent dimensions

To quantify how individual variables contributed to each latent dimension, we computed PLS loadings as the Pearson correlation between the individual-level PLS scores and each original variable. These loadings capture both the strength and direction of association between a variable and a given latent dimension. Higher absolute loadings indicate variables that contribute more strongly to the underlying pattern of covariation, thereby identifying the traits and diagnoses that most prominently shape each PLS dimension. Finally, we employed a cluster bootstrap resampling procedure to evaluate the stability of the PLS loadings. In each of 10,000 iterations, families were resampled with replacement to generate bootstrap samples of the same size as the original dataset, with all siblings from selected families retained together to preserve within-family dependence. The ordering and orientation of PLS dimensions were aligned across iterations to ensure comparability of loadings. This yielded 10,000 bootstrap estimates for each loading, forming an empirical sampling distribution. A loading was deemed robust if its two-sided 95% confidence interval (2.5th to 97.5th percentile) did not include zero, indicating a consistent contribution to the latent dimension across resamples.

#### Time-to-event analysis of ADHD diagnosis

Time-to-event analyses were performed using the *lifelines* Python package (v0.30.0). To examine associations between latent temperament dimensions and the timing of psychiatric diagnosis, we fitted Cox proportional hazards models^68^ using age (in months) as the time scale. For each child, time-to-event was defined as either the age at first recorded diagnosis or the age at last registry entry for those without a diagnosis. Predictors included the PLS scores along with the covariates used in the FEMA-Long models (first twenty genetic principal components). Year of birth and sex were included as stratification variables, allowing the baseline hazard to vary across strata rather than assuming proportionality for these predictors. Models used age-based delayed entry, such that participants entered the risk set at age 5 and were followed until diagnosis or censoring at age 21. Left truncation at age 5 ensured that exposure measures were fully defined before participants entered the risk set. We used cluster-robust variance estimates with family ID as the clustering variable (cluster_col=’fid’, robust=True). Full model outputs are provided in Table S1.

We assessed the proportional hazards assumption using tests and visual inspection of scaled Schoenfeld residuals (implemented in *lifelines*). Sex showed clear evidence of non-proportional hazards for ADHD diagnosis (ICD-10: F90.0) and was therefore treated as a stratification variable, allowing sex-specific baseline hazards. For the PLS dimensions, diagnostics indicated only minor deviations from proportionality. Cox model estimates are thus interpreted as average effects over the follow-up period. Full diagnostics are provided in Fig. S13.

To visualize cumulative diagnosis risk across levels of the temperament-derived dimensions, participants were stratified into quantile-based groups according to their latent dimension scores. Kaplan–Meier curves were then constructed for each group to estimate the survival function, that is, the probability of remaining diagnosis-free up to a given age. Cumulative risk was expressed as *1-S(t)*, corresponding to the proportion of participants diagnosed by age *t*. Furthermore, we examined sex differences by generating separate Kaplan–Meier curves for males and females, without further stratification by PLS score. Together, these plots provide complementary perspectives on how variation in the latent temperament dimensions and sex relate to cumulative diagnosis risk over development.

As a sensitivity analysis, we generated Kaplan–Meier curves using a reference covariate profile. Continuous predictors were fixed at their sample means, categorical predictors at their reference categories and year of birth at the median stratum. Only the latent dimension scores were varied. This allowed us to visualize diagnosis risk for a typical participant and verify that the observed survival differences arose from variation in the latent temperament dimensions rather than covariate heterogeneity (Fig. S14).

A limitation of NPR is that nationwide coverage began in 2008. For children born before 2004, the registry gap potentially overlaps with the early risk period following temperament assessment. For the example of ADHD, twelve children with ADHD diagnosis in the analysis sample were born in 2002–2003. Therefore, their first recorded diagnosis occurred at ages 6–7, but could have occurred earlier without being captured, leading to overestimated time-to-event times. Excluding these twelve children from the time-to-event analysis yielded estimates identical to the main analysis, confirming that this potential bias does not materially affect the reported associations.

#### Identification of shared genetic loci

Given the strong phenotypic association between temperament and ADHD, we examined whether the two phenotypes share genetic architecture. To identify jointly associated loci, we applied conjFDR analysis^37,38^. ConjFDR extends the conditional FDR (condFDR) framework by re-ranking association statistics for a primary phenotype conditional on their association with a secondary phenotype. The re-ranking exploits cross-trait pleiotropy to enhance detection of shared genetic signals. For each pair of traits, condFDR is estimated in both directions, and conjFDR is defined as the maximum of the two corresponding condFDR values, providing a conservative estimate of the probability that a variant is a false positive jointly for both traits. Genomic inflation was controlled using the standard empirical null correction implemented in the conjFDR framework^37,38^. Consistent with prior studies, loci with conjFDR *p* < 0.05 were considered jointly associated. We excluded variants from the major histocompatibility complex (MHC) region, the 8p23.1 locus, and the 17q21.31 inversion region to limit inflation driven by the extended linkage disequilibrium structure characteristic of these regions.

As input to conjFDR analyses, we used summary statistics from genome-wide association studies. ADHD summary statistics were obtained from the Psychiatric Genomics Consortium^69^ (PGC; 38,691 cases and 186,843 controls). For temperament, we used GWAS summary statistics derived from FEMA-Long, including both main genetic effects and age-dependent genetic effects.

To identify lead SNPs in each conjFDR analysis, we performed LD-based clumping using a threshold of *r*² < 0.6 to obtain a set of approximately independent index variants. For each index SNP, all variants in high linkage disequilibrium (*r*² ≥ 0.6) were identified using the 1000 Genomes Project European reference panel. Loci located within 250 kb of each other were subsequently merged, and within each merged locus, the SNP with the lowest conjFDR value was selected as the lead SNP.

#### Gene mapping and phenotypic annotation

To functionally characterize genetic loci identified in the conjFDR analyses, we annotated candidate SNPs using the SNP2GENE pipeline implemented in FUMA^70^. Candidate SNPs were defined as all variants reaching conjFDR *p* < 0.05, including independent lead SNPs as well as all SNPs in linkage disequilibrium with these lead variants (*r*² ≥ 0.6). SNP-to-gene mapping was performed using the default FUMA settings based on positional mapping. To contextualize the resulting genes, we queried their previously reported trait associations in the GWAS Catalog^39^. For each mapped gene, we extracted the number of reported associations per gene and the frequency of each trait across genes.

## Acknowledgements

The Norwegian Mother, Father and Child Cohort Study is supported by the Norwegian Ministry of Health and Care Services and the Ministry of Education and Research. We are grateful to all the participating families in Norway who take part in this on-going cohort study. For generating high-quality genomic data, we thank the Norwegian Institute of Public Health (NIPH), the HARVEST collaboration, the NORMENT Centre at the University of Oslo, the Center for Diabetes Research at the University of Bergen, deCODE Genetics, the Research Council of Norway, the South-Eastern and Western Norway Regional Health Authorities, the ERC AdG, Stiftelsen KG Jebsen, the Trond Mohn Foundation, and the Novo Nordisk Foundation.

This work was partly performed on the TSD (Tjeneste for Sensitive Data) facilities, owned by the University of Oslo, operated and developed by the TSD service group at the University of Oslo, IT-Department (USIT). Computations were also performed on resources provided by UNINETT Sigma2 - the National Infrastructure for High-Performance Computing and Data Storage in Norway (NS9703S).

The authors were funded by the Research Council of Norway (296030, 324252, 324499, 326813, 334920, 351751); the South-Eastern Norway Regional Health Authority (2020060 (IES), 2025037 (IES)); European Union’s Horizon 2020 Research and Innovation Programme (Grant No. 847776; CoMorMent and Grant No. 964874; RealMent); EU’s Horizon Psych-STRATA project (#101057454); Stiftelsen KG Jebsen (SKGJ-MED-021), National Institutes of Health (NIH; U24DA041123; R01AG076838; U24DA055330; and OT2HL161847). JK was supported by a Marie Skłodowska-Curie Postdoctoral Fellowship under the European Union’s Horizon Europe research and innovation programme (Grant Agreement No. 101150746). EPT was supported by a Rubicon Postdoctoral Fellowship of ZonMW (Grant No. 04520242420017). PP is supported by the National Institutes of Health grants U24DA041123 and U24DA055330, and by Wellcome Leap, CARE Program (“FEMA-AD”).

## Data availability

Data from the Norwegian Mother, Father and Child Cohort Study (MoBa) and the Medical Birth Registry of Norway are administered by the Norwegian Institute of Public Health and are not publicly available due to ethical and legal restrictions. Access may be granted to qualified researchers following approval from the Regional Committees for Medical and Health Research Ethics (REC), compliance with the EU General Data Protection Regulation (GDPR), and permission from the data owners. Participant consent does not allow for deposition of individual-level data in public repositories. Researchers seeking access for replication or further analyses must apply through the Norwegian health data access portal (helsedata.no) and enter a data access agreement with MoBa.

## Code availability

FEMA-Long is available on GitHub at: https://github.com/cmig-research-group/cmig_tools. conjFDR is available on GitHub at: https://github.com/precimed/pleiofdr.

Custom analysis software used in this work is available in a publicly accessible GitHub repository: https://github.com/jakubkopal/temperamet_deviations

## Conflicts of interest

OAA has received speaker’s honorarium from Lundbeck, Janssen, Otsuka, and Sunovion and is a consultant to Cortechs.ai. and Precision Health. AMD is Founding Director, holds equity in CorTechs Labs, Inc. (DBA Cortechs.ai), and serves on its Board of Directors. AMD is the President of J. Craig Venter Institute (JCVI) and is a member of the Board of Trustees of JCVI. He is an unpaid consultant for Oslo University Hospital. OF is a consultant to Precision Health. All other authors report no potential conflicts of interest.

## Notes

### Author Declarations

The establishment and data collection of MoBa were approved by the Norwegian Data Protection Agency and the Regional Committees for Medical and Health Research Ethics (REK). MoBa operates under the Norwegian Health Registry Act. The current study was approved by the MoBa administrative board at the Norwegian Institute of Public Health (NIPH) and REK (reference 2016/1226/REK Sor-Ost C).

### Summary of Updates

Author name spelling corrected. No changes to text, figures, or analyses.

