## Supplementary Materials for "Early-childhood temperament deviations mark psychiatric risk into early adulthood"

### Supplementary Figures

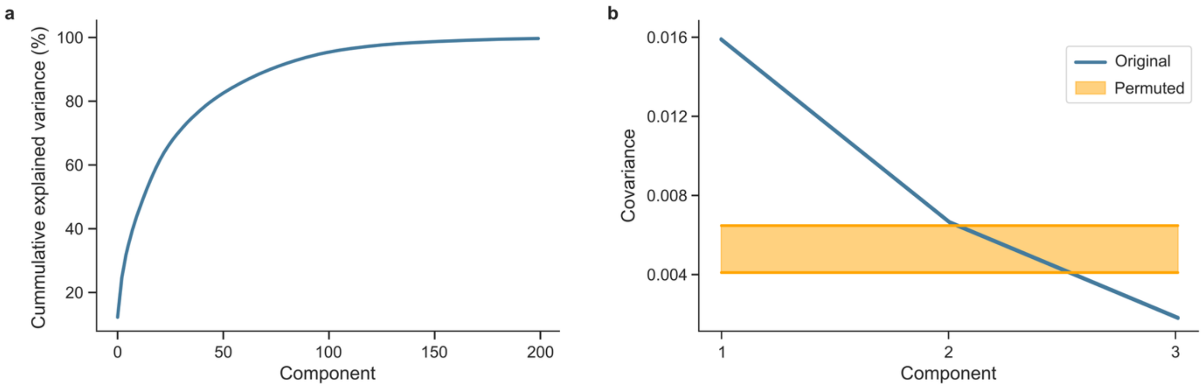


**Figure S1: Selected number of PCA components and PLS dimensions**

**a.** Number of PCA components. Cumulative explained variance for successive principal components derived from the binary ICD-10 diagnostic matrix are shown. The first 100 components collectively accounted for 95% of the total variance. **b.** Number of PLS dimensions. To determine the number of significant PLS dimensions linking temperament and diagnoses, we performed a permutation-based significance test. For each PLS mode, the observed covariance between temperament and diagnostic scores was compared against a null distribution generated from 10,000 permutations. In each iteration, the alignment between temperament deviations and diagnostic measures was disrupted by permuting participants at the family level (i.e., shuffling family blocks while keeping siblings together), thereby preserving within-family dependence while breaking cross-dataset correspondence. The resulting null distribution for the first PLS mode is shown in yellow. P-values were computed as the proportion of permuted covariances that exceeded the observed covariance for each mode. The first two PLS dimensions had $p$-values $< 1\times10^{-5}$.

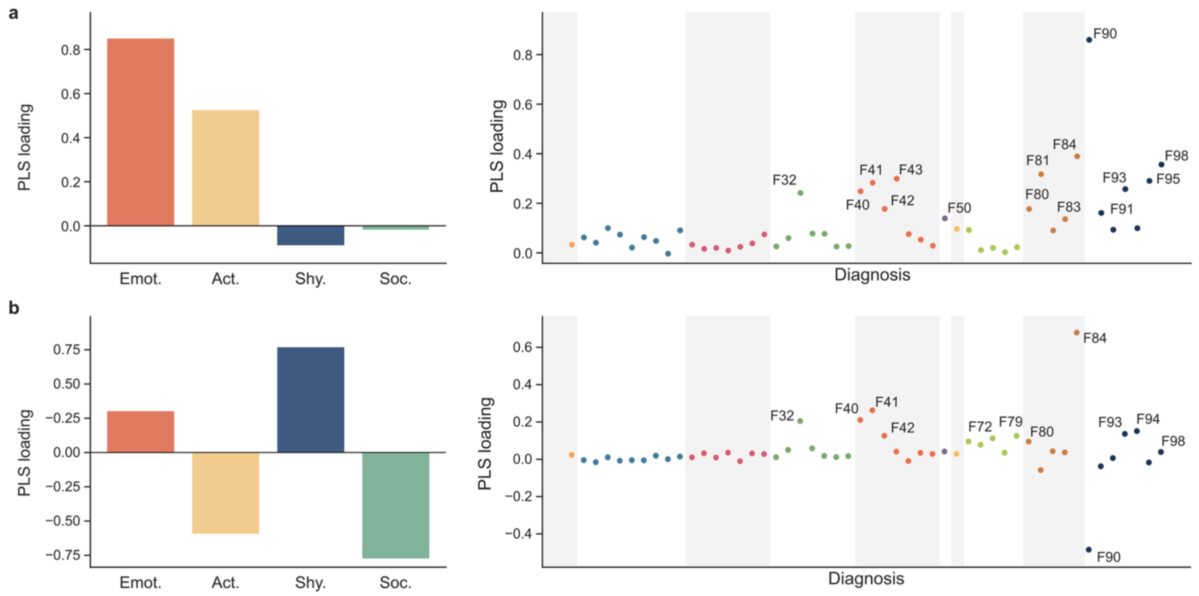


**Figure S2: Latent dimensions of temperament deviations and diagnostic categories**

To assess the robustness of the presented PLS analysis, we repeated the analysis using ICD-10 category codes, which aggregate the 266 subcategory codes into 50 broader diagnostic categories. Rerunning the PLS model yielded again two significant dimensions. The plot depicts the first (**a**) and second (**b**) PLS dimensions. Bar plots show the temperament loadings, while dot plots display the diagnostic loadings. The temperament loadings closely resemble those reported in the main analysis. Similarly, the diagnostic loadings mirror the more detailed results presented in the main article. Consistent with the primary analysis, hyperkinetic disorders (F90), which include ADHD diagnoses, emerged as the most prominent diagnostic category in both dimensions.

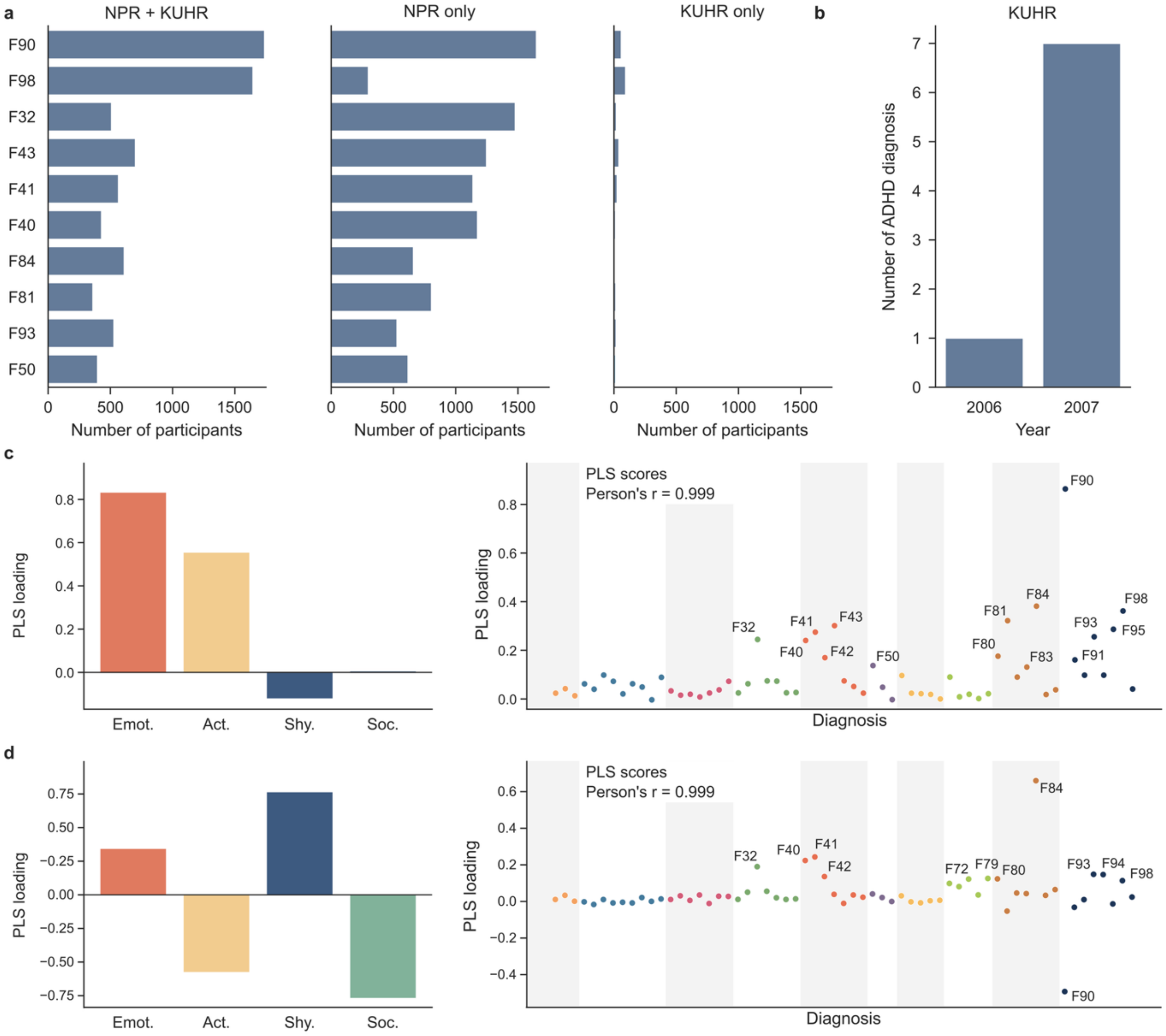


**Figure S3: Multivariate analyses with KUHR as an additional diagnostic source**

KUHR contains reimbursement records for healthcare encounters funded through the Norwegian National Insurance Scheme, including primary care consultations and some specialist services not captured by NPR. Importantly, KUHR records date back to 2006, compared to 2008 for NPR. **a.** KUHR and NPR diagnoses comparisons. Bar plot showing the number of lifetime diagnoses present in both NPR and KUHR, diagnoses present only in NPR, and diagnoses present only in KUHR. The ten most common diagnoses based on NPR are shown. **b.** Early KUHR records. ADHD diagnoses recorded in KUHR during the two years not covered by NPR. Because only one ADHD diagnosis was recorded in 2006, there are one or fewer ADHD diagnoses prior to 2006. We then repeated the PLS analysis using ICD-10 category codes derived from both NPR and KUHR. The first (**c**) and second (**d**) PLS dimensions closely mirror those obtained using NPR diagnoses only. Specifically, the temperament PLS loadings (left bar plots) are identical. Diagnostic PLS loadings include 11 additional categories. However, the strongest loadings remain the same as in the NPR-only analysis. In summary, KUHR contributes only a small number of additional diagnoses, resulting in results that are highly similar to those presented in the main analysis.

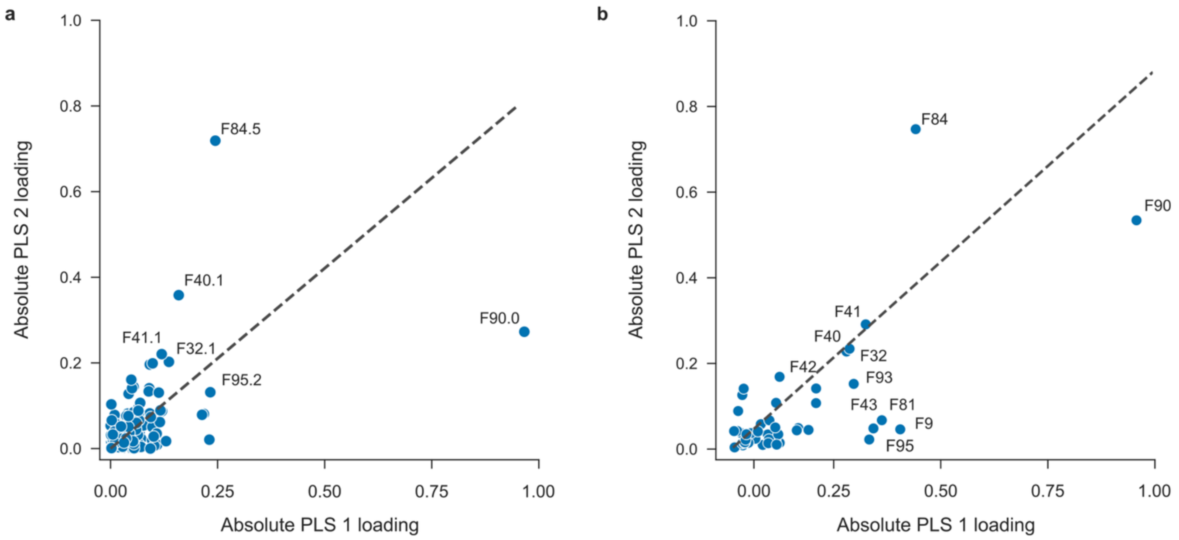


**Figure S4: Diagnoses dominating PLS decomposition**

Comparison of PLS loading magnitudes for the first and second dimensions. Absolute PLS loadings for the first dimension are plotted against those for the second dimension for models using subcategory ICD-10 codes (**a**) and category ICD-10 code groupings (**b**). The dotted line represents the diagonal where loadings are equal across dimensions. ADHD, and its broader diagnostic category of hyperkinetic disorders, show strong contributions to both latent dimensions.

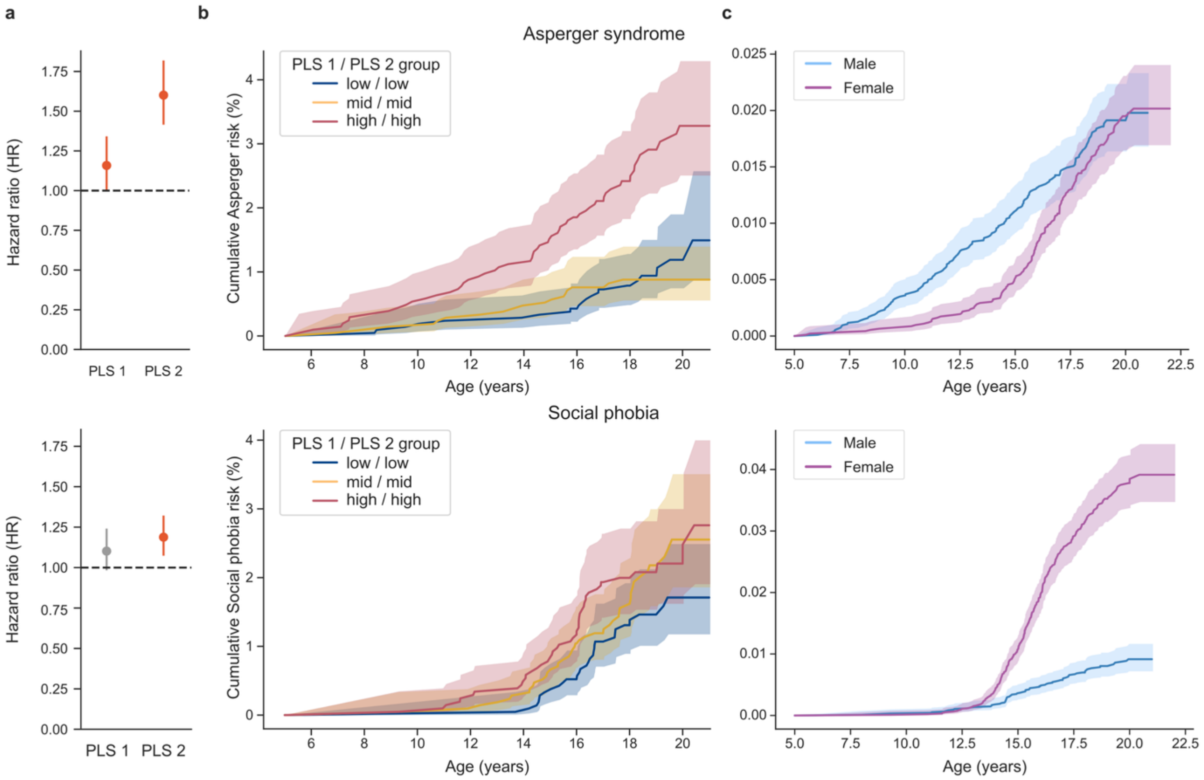


**Figure S5: Risk and timing of Asperger syndrome and social phobia**

We estimated two time-to-event models using the first two PLS dimensions to predict time to diagnosis of Asperger syndrome (top row) and social phobia (bottom row), as these diagnoses loaded most strongly on the second PLS dimension. **a.** Risks across latent temperament dimensions. Hazard ratios from the time-to-event models are shown with 95% confidence intervals. Estimates greater than 1 (risk factors) are shown in red, and estimates less than 1 (protective effects) are shown in blue. **b.** Risk stratification based on combined latent temperament dimensions. Kaplan–Meier curves display cumulative risk of Asperger syndrome and social phobia for three participant groups stratified by joint scores on the first and second PLS dimensions. Curves are shown for groups with low, intermediate, and high combined scores. **c.** Sex-specific disease risk. Kaplan–Meier curves display the cumulative probability of receiving a diagnosis separately for males and females, reflecting sex-specific differences in disease risk across development. Together, latent dimensions capture variation in disease risk and are associated with differences in the timing of diagnosis.

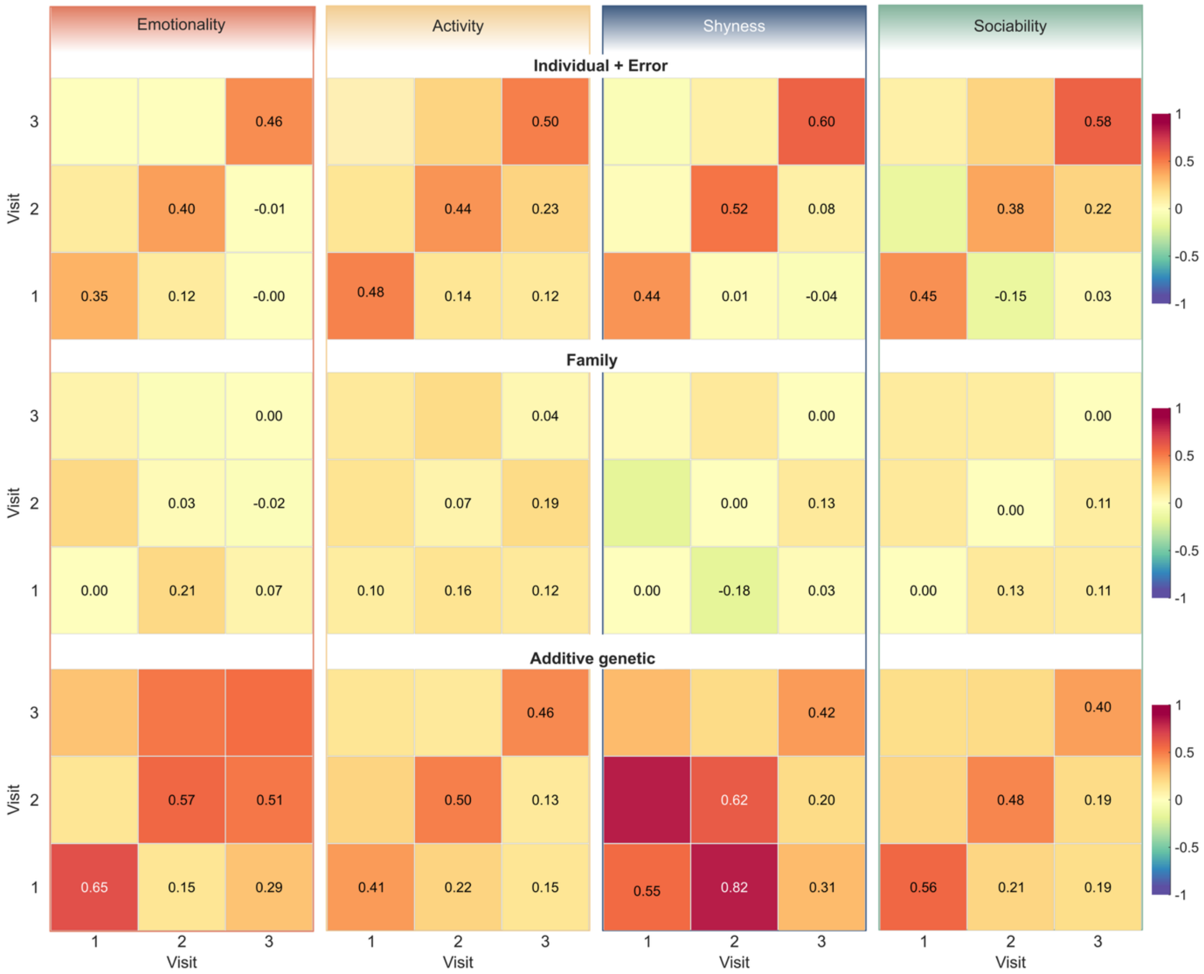


**Figure S6: Variance–covariance structure of temperament across development**
FEMA-Long decomposes phenotypic variability into three sources: individual-level (which also includes noise), family-level, and additive genome-wide genetic variance. For each temperament trait (*emotionality*, *activity*, *shyness*, *sociability*), heatmaps display the estimated variance at each assessment wave (diagonal cells) and the normalized unstructured covariance between timepoints (off-diagonal cells), reflecting temporal stability within each variance component. Warmer colors indicate larger variance or stronger covariance. Together, these matrices illustrate how the magnitude and stability of each source of variation change across ages 1.5, 3, and 5 years.

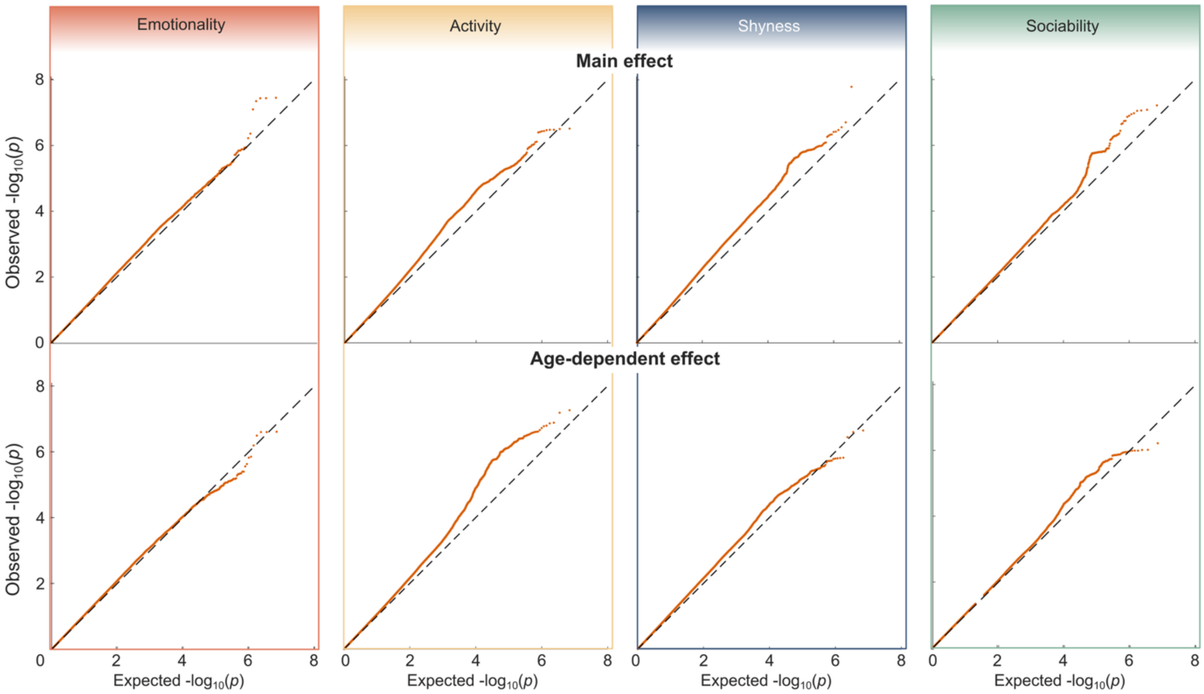


**Figure S7: Q-Q plots of genetic enrichment for main and age-dependent effects**

Q-Q plots compare the observed ($y$-axis) and expected ($x$-axis) $-\log_{10}(p)$ values to assess enrichment of genetic associations. The first row shows results for main genetic effects estimated with FEMA-Long, and the second row shows age-dependent effects. The four columns correspond to the four temperament traits. The diagonal line represents the expected distribution under the null hypothesis. Deviations above the diagonal indicate enrichment, where observed values are more significant than expected, reflecting potential association signals.

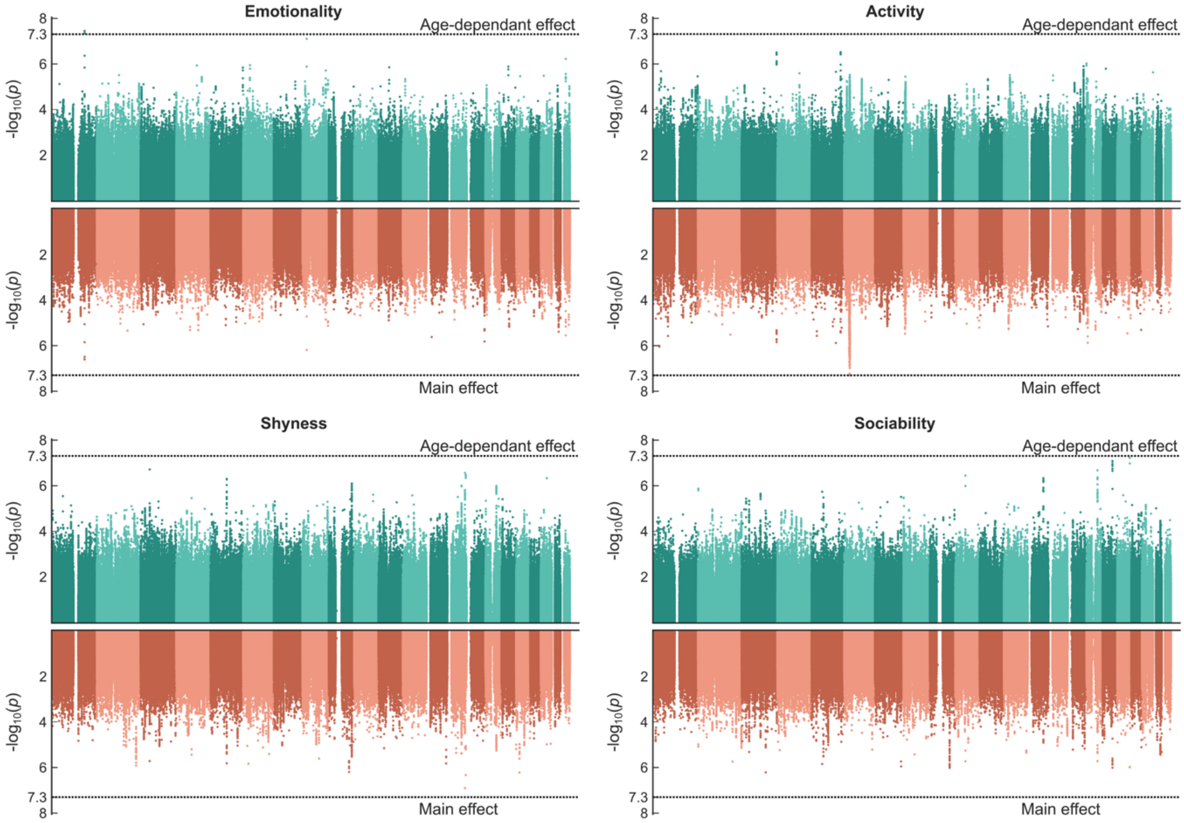


**Figure S8: Miami plots of stable and age-dependent genetic effects**

Miami plots display $-\log_{10}(p)$ values for genome-wide main effects (bottom panels) and age-dependent effects (upper panels) for each temperament trait (*emotionality*, *activity*, *shyness*, *sociability*). The bottom panels reflect SNP effects that remain consistent across development. The upper panels show results from a Wald test evaluating whether the spline-based age effect differs from zero, indicating genetic influences that change with age. Highlighted alternating shading marks chromosome boundaries. The horizontal dashed lines indicate the genome-wide significance threshold ($p < 5\times10^{-8}$). Together, these plots provide a visual comparison of stable versus developmentally dynamic genetic architecture across temperament traits.

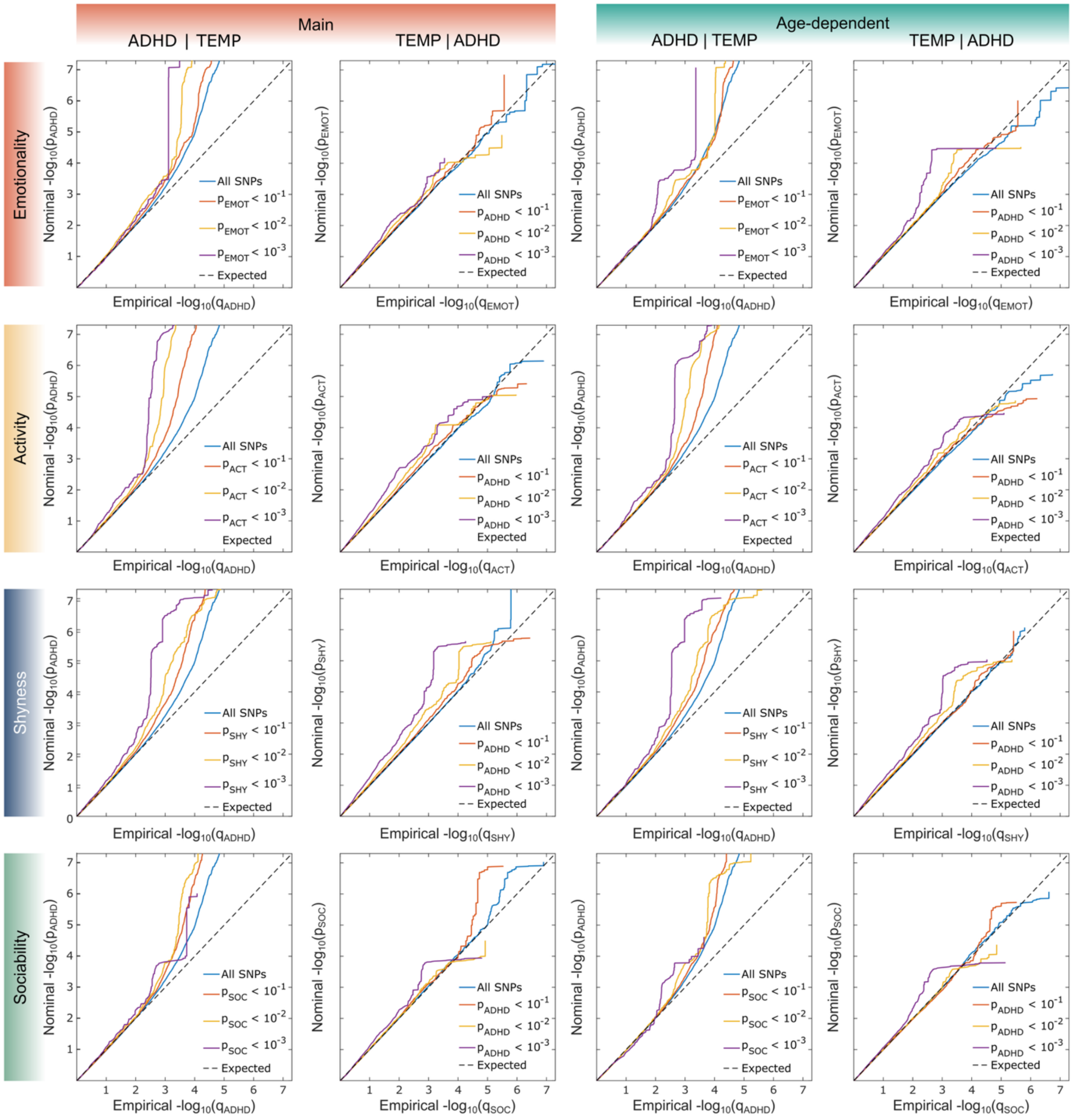


**Figure S9: Q-Q plots of genetic enrichment for shared loci**

We performed conjunctional false discovery rate (conjFDR) analyses to identify genetic loci jointly associated with temperament traits and ADHD. The genetic architecture of temperament was represented using SNP-level significance for both main genetic effects and age-dependent effects estimated with FEMA-Long. The Q–Q plots compare observed $-\log_{10}(p)$ values ($y$-axis) to those expected under the null hypothesis ($x$-axis) to assess cross-trait enrichment of genetic associations, a key prerequisite for applying the conjFDR framework. The diagonal line indicates the null expectation. Colored curves represent SNP subsets stratified by increasing association strength in the conditioning trait. The first and third columns show enrichment of ADHD conditional on the temperament trait, whereas the second and fourth columns show enrichment of the temperament trait conditional on ADHD. Systematic deviations above the diagonal indicate enrichment, reflecting an excess of shared genetic associations beyond chance.

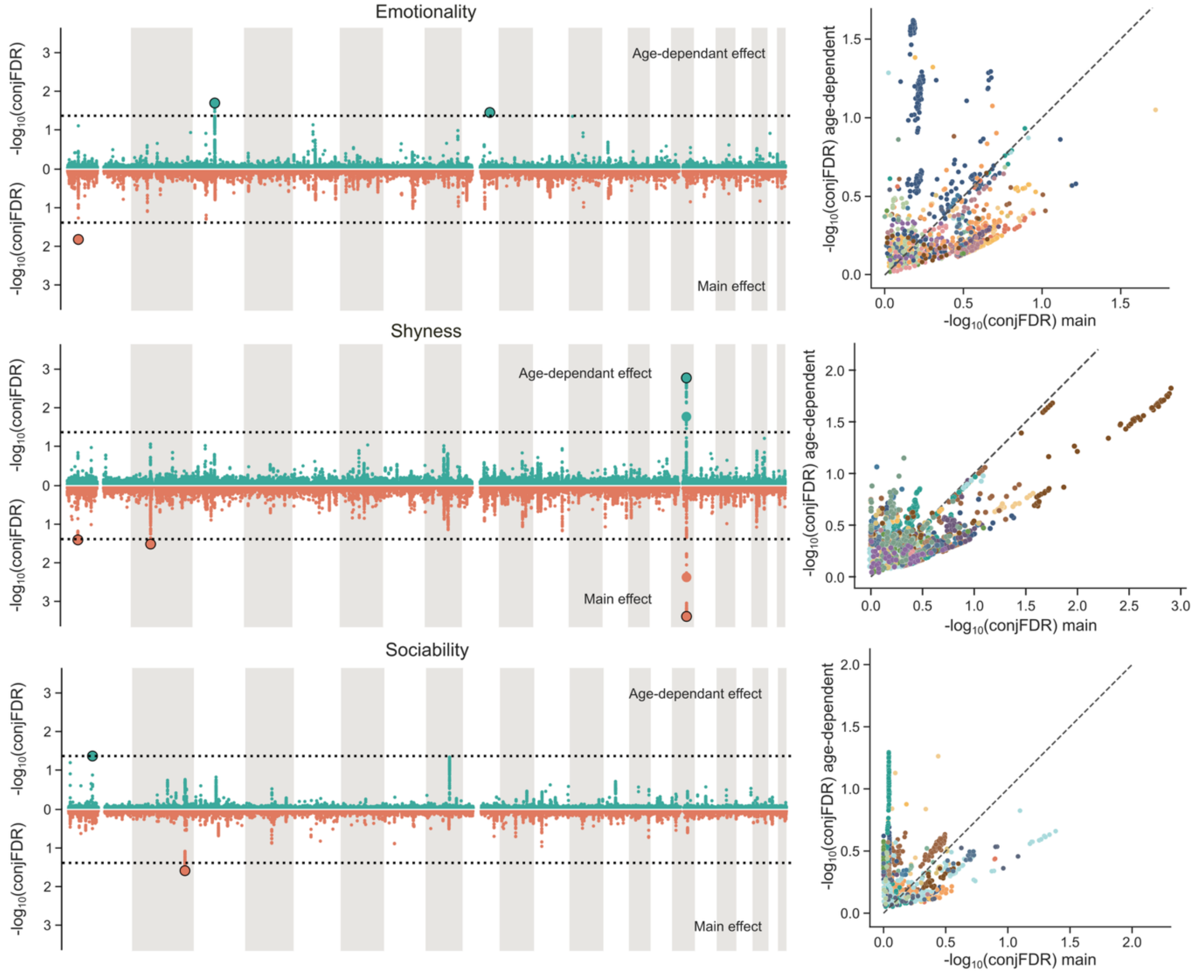


**Figure S10: Miami plots of main and age-dependent genetic effects identified by conjFDR** To complement the activity results shown in Fig. 5c, we present the corresponding conjFDR results for *emotionality*, *shyness*, and *sociability*. **Left:** Joint genetic associations between each temperament trait and ADHD. Miami plots display SNPs jointly associated with ADHD and the respective temperament trait, shown separately for the main genetic effect (bottom) and the age-dependent genetic effect (top). The $y$-axis shows $-\log_{10}$-transformed conjFDR values. Black circles denote significant lead SNPs within linkage disequilibrium blocks. **Right:** Genome-wide comparison of association strength for main and age-dependent effects. The plot shows $-\log_{10}$-transformed conjFDR values for the main genetic effects against the age-dependent genetic effects across all SNPs. Colors denote different chromosomes.

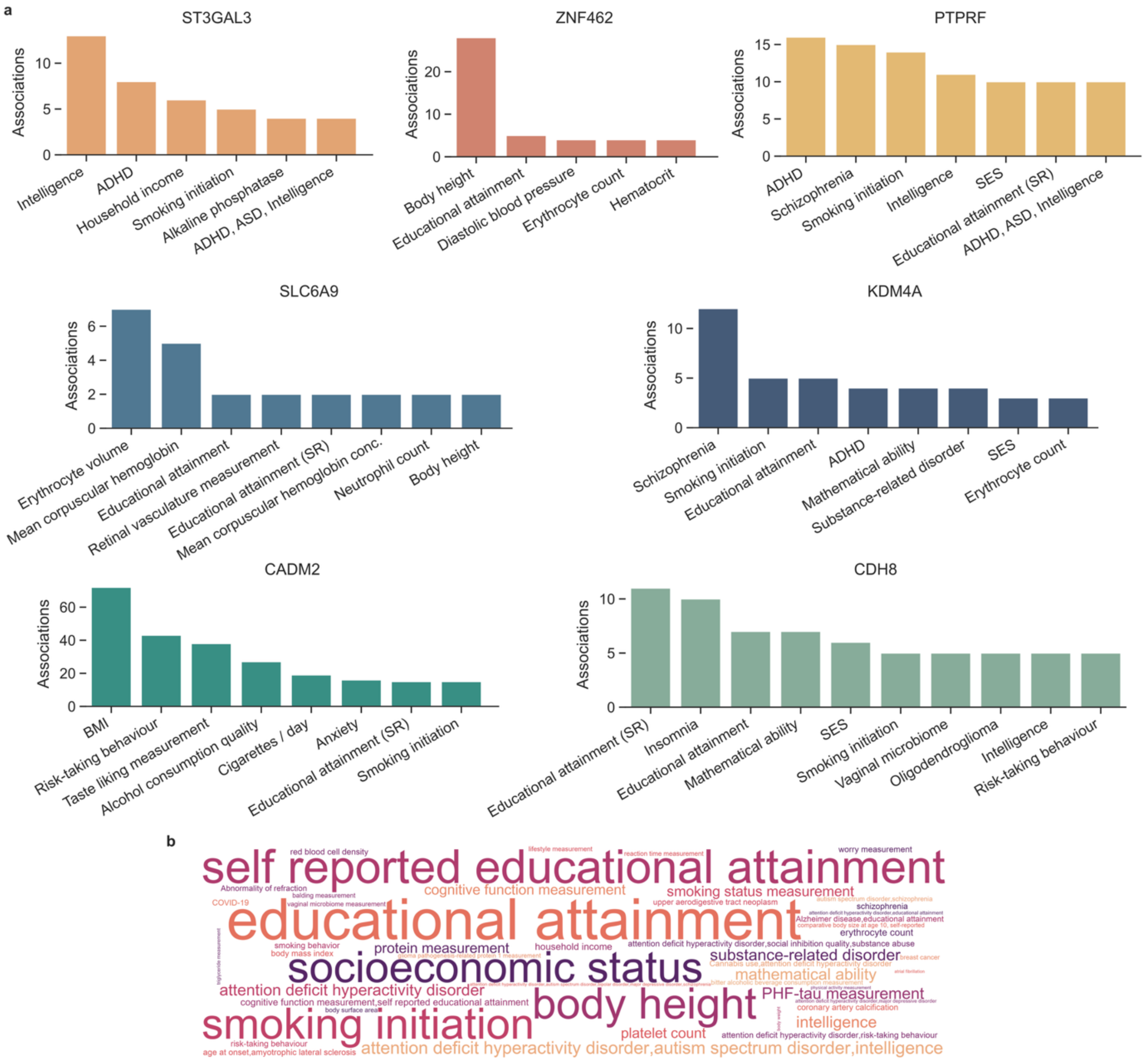


**Figure S11: Previously reported trait associations for temperament-implicated genes**
Lead SNPs identified through conjFDR analysis were mapped to genes based on genomic position (FUMA). The resulting genes were queried against the GWAS Catalog to identify previously reported trait associations. **a.** Association counts per gene. Bar plots show the number of GWAS Catalog entries linking each gene to specific traits. The *x*-axis lists previously reported traits. The *y*-axis shows the count of catalog entries for each gene-trait pair. **b.** Trait frequency across genes. Word cloud displays how many genes have been previously associated with each trait. The word size corresponds to the gene count. Educational attainment was reported for seven mapped genes, while body height, socioeconomic status, smoking initiation, ADHD, autism spectrum disorder, and intelligence were each reported for six of the eight genes. Abbreviations: ADHD, attention-deficit/hyperactivity disorder; ASD, autism spectrum disorder; SES, socioeconomic status; BMI, body mass index; SR, self-reported.

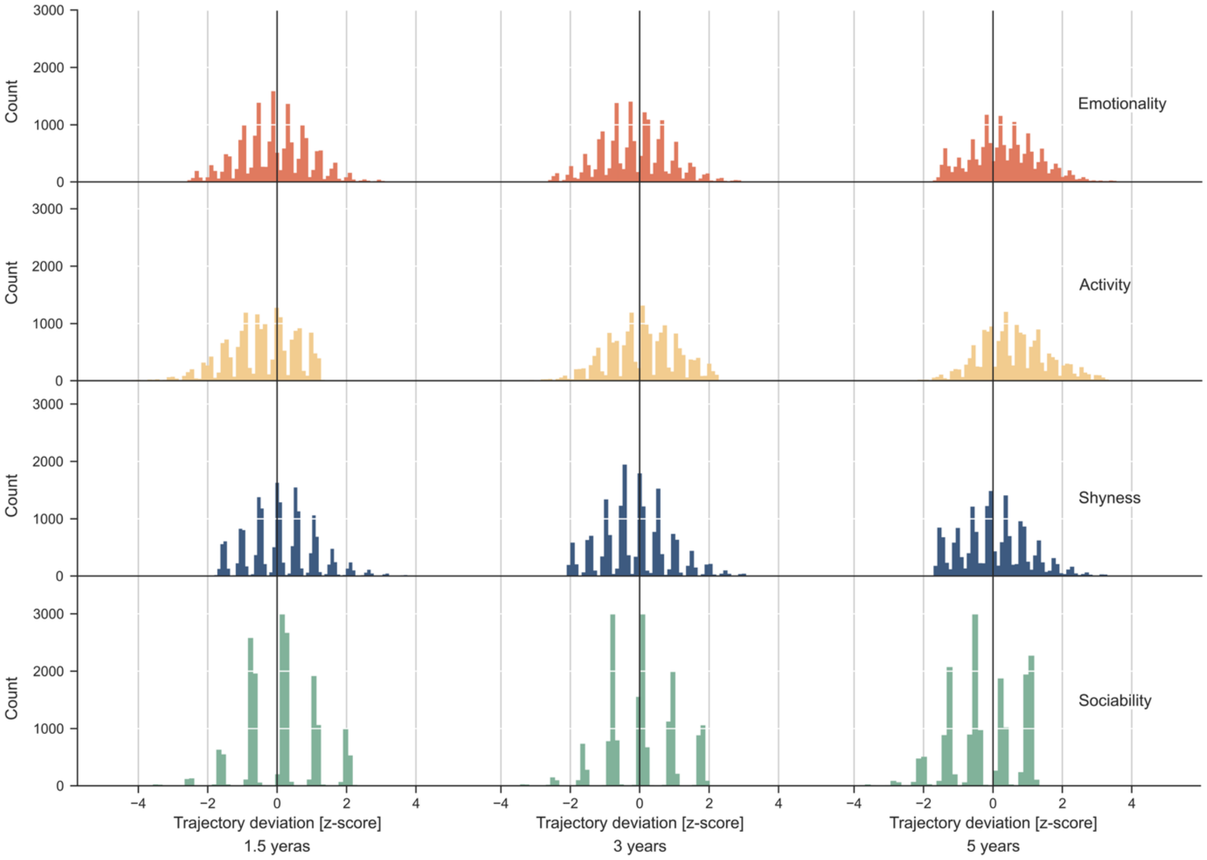


**Figure S12: Distribution of deviations from predicted temperament trajectories**
Temperament deviations were calculated for 18,651 children with complete data across all three timepoints by subtracting each child's observed standardized temperament score from the corresponding fixed effects prediction. Histograms show the distribution of these deviations for each temperament trait (*emotionality*, *activity*, *shyness*, *sociability*) at ages 1.5, 3, and 5 years. The vertical line at zero represents perfect alignment with the predicted developmental trajectory. Positive and negative values indicate children scoring above or below their expected level at that age. These distributions summarize the inter-individual variability in how children deviate from normative temperament development.

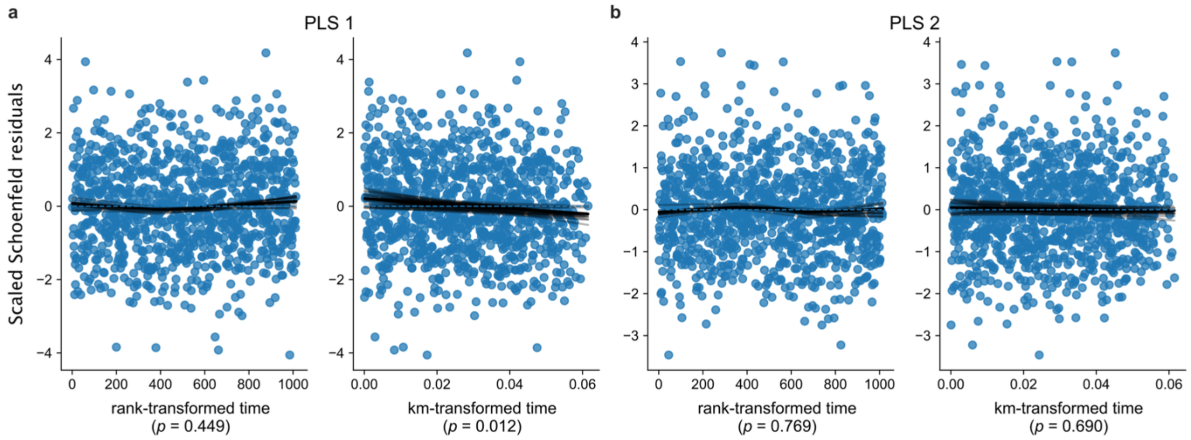


**Figure S13: Assessment of the proportional hazards assumption**

Scaled Schoenfeld residuals for the first (**a**) and second (**b**) latent temperament dimensions from the Cox proportional hazards model predicting ADHD diagnosis. Residuals are plotted against rank-transformed time (left) and Kaplan–Meier–transformed time (right), with locally smoothed trends (LOWESS, solid black lines) overlaid. Non-horizontal trends indicate potential violations of the proportional hazards assumption. P-values from tests based on scaled Schoenfeld residuals are shown below each panel. For the second PLS dimension, no evidence of non-proportional hazards was observed (both $p > 0.68$). For the first PLS dimension, the rank-transformed diagnostic showed no violation ($p$ = 0.45), whereas the Kaplan–Meier–transformed diagnostic indicated evidence of time dependence ($p$ = 0.01). Visual inspection of the LOWESS trends suggested no substantial systematic deviation from horizontality.

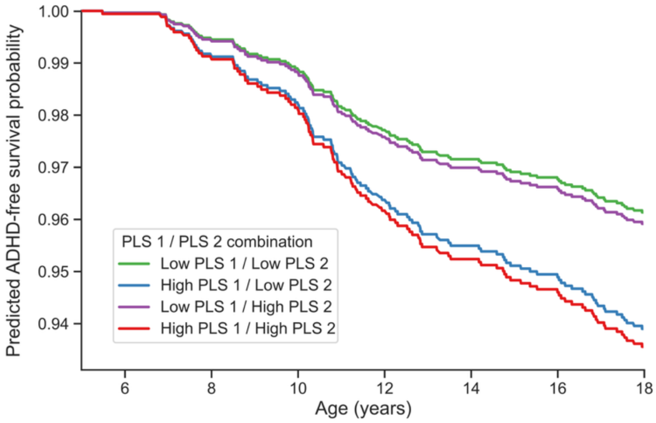


**Figure S14: ADHD-free survival stratified by latent temperament dimensions**

To assess whether temperament-derived latent dimensions meaningfully stratify ADHD risk, we generated Kaplan–Meier survival curves based on the first and second PLS dimensions. Participants were grouped into four strata based on high versus low scores on each dimension. Curves reflect predicted ADHD-free survival using a reference covariate profile, where continuous predictors were set to their sample means, categorical predictors to reference categories, and birth year to the median stratum, allowing only the two PLS dimensions to vary. The observed curve separation indicates that the combination of latent PLS dimensions differentiates ADHD risk independent of other covariates.

### Supplementary Tables

**Table S1: Cox models relating latent temperament dimensions to ADHD diagnosis risk**
The table reports hazard ratios (HRs), 95% confidence intervals (CIs), and *p* values for associations between covariates and time to first ADHD diagnosis during follow-up estimated using Cox proportional hazards model.

| Covariate | Hazard ratio | 95% CI (lower) | 95% CI (upper) | *p*-value |
|---|---|---|---|---|
| **PLS 1** | 1.524 | 1.410 | 1.647 | < 0.001 |
| **PLS 2** | 1.046 | 0.979 | 1.117 | 0.183 |
| **Genetic PC 1** | 1.117 | 1.052 | 1.185 | < 0.001 |
| **Genetic PC 2** | 0.982 | 0.925 | 1.042 | 0.539 |
| **Genetic PC 3** | 0.996 | 0.940 | 1.055 | 0.889 |
| **Genetic PC 4** | 1.039 | 0.978 | 1.104 | 0.212 |
| **Genetic PC 5** | 1.051 | 0.985 | 1.121 | 0.134 |
| **Genetic PC 6** | 1.008 | 0.952 | 1.068 | 0.778 |
| **Genetic PC 7** | 1.001 | 0.940 | 1.066 | 0.983 |
| **Genetic PC 8** | 0.974 | 0.918 | 1.033 | 0.384 |
| **Genetic PC 9** | 0.946 | 0.892 | 1.003 | 0.063 |
| **Genetic PC 10** | 1.016 | 0.962 | 1.074 | 0.566 |
| **Genetic PC 11** | 1.059 | 0.996 | 1.126 | 0.069 |
| **Genetic PC 12** | 0.964 | 0.908 | 1.025 | 0.242 |
| **Genetic PC 13** | 1.068 | 1.006 | 1.135 | 0.031 |
| **Genetic PC 14** | 0.961 | 0.909 | 1.017 | 0.170 |
| **Genetic PC 15** | 1.042 | 0.985 | 1.102 | 0.152 |
| **Genetic PC 16** | 0.936 | 0.884 | 0.991 | 0.023 |
| **Genetic PC 17** | 0.966 | 0.908 | 1.027 | 0.263 |
| **Genetic PC 18** | 1.023 | 0.962 | 1.088 | 0.461 |
| **Genetic PC 19** | 1.016 | 0.956 | 1.080 | 0.605 |
| **Genetic PC 20** | 0.972 | 0.916 | 1.030 | 0.335 |

**Table S2: Lead SNPs identified by conjFDR analysis and mapped genes**
The table lists lead SNPs reaching conjFDR significance and their mapped genes based on FUMA annotation. Gene identifiers are provided using Ensembl nomenclature.

| Ensembl gene ID | Gene symbol | Chromosome | Lead SNPs |
|---|---|---|---|
| ***Emotionality* main & ADHD** | | | |
| ENSG00000126091 | ST3GAL3 | 1 | rs113551349 |
| ENSG00000196517 | SLC6A9 | 1 | rs113551349 |
| ***Emotionality* age-dependent effect & ADHD** | | | |
| ENSG00000175161 | CADM2 | 3 | rs12637461 |
| ENSG00000148143 | ZNF462 | 9 | rs149916490 |
| ENSG00000242631 | RP11-508N12.4 | 9 | rs149916490 |
| ***Activity* main effect & ADHD** | | | |
| ENSG00000142949 | PTPRF | 1 | rs10789442 |
| ENSG00000066135 | KDM4A | 1 | rs10789442 |
| ENSG00000126091 | ST3GAL3 | 1 | rs10789442 |
| | | 6 | rs1343667 |
| ***Activity* age-dependent effect & ADHD** | | | |
| ENSG00000142949 | PTPRF | 1 | rs10789442 |
| ENSG00000066135 | KDM4A | 1 | rs10789442 |
| ENSG00000126091 | ST3GAL3 | 1 | rs10789442 |
| | | 6 | rs1343667 |
| | | 11 | rs6484367 |
| ***Shyness* main effect & ADHD** | | | |
| ENSG00000142949 | PTPRF | 1 | rs489319 |
| ENSG00000066135 | KDM4A | 1 | rs489319 |
| ENSG00000126091 | ST3GAL3 | 1 | rs489319 |
| | | 2 | rs7570163 |
| ENSG00000150394 | CDH8 | 16 | rs8044799 |
| ***Shyness* age-dependent effect & ADHD** | | | |
| ENSG00000150394 | CDH8 | 16 | rs1397128 |
